# High Dimensional Mediation Analysis: a new method applied to maternal smoking, placental DNA methylation and birth outcomes

**DOI:** 10.1101/2022.03.15.22272404

**Authors:** Basile Jumentier, Claire-Cécile Barrot, Maxime Estavoyer, Jorg Tost, Barbara Heude, Olivier François, Johanna Lepeule

## Abstract

**Background:** High-dimensional mediation analysis is an extension of unidimensional mediation analysis that includes multiple mediators, and is increasingly used to evaluate the indirect omics-layer effects of environmental exposures on health outcomes. Analyses involving high-dimensional mediators raise several statistical issues. While many methods have recently been developed, no consensus has been reached about the optimal combination of approaches to high-dimensional mediation analyses.

**Objectives:** We developed and validated a method for high-dimensional mediation analysis (HDMAX2) and applied it to evaluate the causal role of placental DNA methylation in the pathway between exposure to maternal smoking (MS) during pregnancy and gestational age (GA) and weight (BW) of the baby at birth.

**Methods:** HDMAX2 combines latent factor regression models for epigenome-wide association studies with max-squared tests for mediation, and considers CpGs and aggregated mediator regions (AMR). HDMAX2 was carefully evaluated on simulated data, and compared to state-of-the-art multi-dimensional epigenetic mediation methods. Then HDMAX2 was applied on data from 470 women of the EDEN cohort.

**Results:** HDMAX2 demonstrated increased power compared to state-of-the-art multi-dimensional mediation methods, and identified several AMRs not identified in previous mediation analyses of exposure to MS on BW and GA. The results provided evidence for a polygenic architecture of the mediation pathway with an overall indirect effect of CpGs and AMRs of 44.5 g lower BW (32.1% of the total effect). HDMAX2 also identified AMRs having simultaneous effects both on GA and on BW. Among the top hits of both GA and BW analyses, regions located in *COASY, BLCAP* and *ESRP2* also mediated the relationship between GA on BW, suggesting a reverse causality in the relationship between GA and the methylome.

**Discussion:** HDMAX2 outperformed existing approaches and revealed an unsuspected complexity of the causal relationships between exposure to MS and BW at the epigenome-wide level. HDMAX2 is applicable to a wide range of tissues and omic layers.

## 1. Introduction

Mediation analysis is a statistical tool used to gain insights into the causal mechanisms that relate an exposure to an outcome (1). It is increasingly used in environmental epidemiology, in particular in Developmental Origins of Health and Disease (DOHaD) research and in molecular epidemiology studies (2,3). With the development of high-throughput screening technologies, these methods have become key tools to investigate the pathways by which environmental exposures can affect health outcomes, and more specifically those involving epigenetic mechanisms such as DNA methylation (DNAm) variations (4–7).

High-dimensional mediation analysis is an extension of unidimensional mediation analysis including multiple mediators (2). A typical high-dimensional analysis for DNAm markers generally includes three main steps. The first step tests both the effects of exposure on DNAm levels and the effects of DNAm levels on the health outcome based on epigenome-wide association studies (EWAS). The second step combines significance values obtained from the two EWAS at the first step in order to perform mediation tests, and assesses the mediator status of each marker. The third step quantifies the indirect effects of exposure on the health outcome through DNAm differences. Analyses involving a large set of mediators are difficult, and raise numerous statistical issues (2). To overcome those issues, several approaches have been proposed during the recent years. Classical approaches perform multi-dimensional analysis by running unidimensional mediation analyses for each DNAm marker, for example using Sobel tests or by estimating Average Causal Mediated Effects (ACME) (8,9). Improvements of the Sobel test for indirect effects combine the significance values obtained from the two EWAS in various ways (10–15). However, there is no consensus on the most relevant combination of EWAS and mediation tests for a high-dimensional analysis. Furthermore, the overall indirect effect of multiple mediators remains poorly quantified from estimates of single mediator effects in a context of correlation among the mediators.

We addressed the above issues by developing HDMAX2, a method for high-dimensional mediation analysis, and systematically compared HDMAX2 to recently proposed approaches. HDMAX2 relies on latent factor regression models to evaluate associations of exposure and outcome with DNAm, and on mediation tests that control the type I error when combining the significance values obtained in the exposure and outcome EWAS. We developed additional features to further consider methylation regions as mediators and to estimate an overall mediated effect of DNAm accounting for all identified mediators simultaneously. We then used HDMAX2 to evaluate the causal role of placental DNA methylation in the pathway between maternal smoking during pregnancy, gestational age at delivery and birth weight of the baby. Several studies focused on cord blood (16–19). Although it plays a key role in fetal programming, few studies have investigated placental DNAm (20–23). We discovered new CpG mediators and regions that were not identified by classic approaches (Cardenas et al., 2019; Morales et al., 2016), and we estimated the overall indirect effect of maternal smoking during pregnancy on newborn birth weight and on gestational age at delivery. The approaches we used for placental DNAm data also holds for other types of tissue or quantitative omics data, and HDMAX2 can extend to various types of data.

## 2. Methods

### 2.1 Overview of the HDMAX2 method

HDMAX2 is a new approach for high dimensional mediation analysis structured in three main steps (Figure 1). The first step of HDMAX2 corresponds to an extension of regression models considered generally in unidimensional mediation analysis (1). The extension includes latent factors as covariates in the models to account for unobserved variables that confound multidimensional DNAm data analysis, such as batch effects or cell-type heterogeneity in samples. The second step identifies potential mediators by combining paired significance values that are obtained when testing the effect of exposure on DNAm and the effect of DNAm on outcome in step 1. Step 2 is not restricted to CpG markers, and it can also identify aggregated mediator regions (AMRs) based on the paired *P*-values. The third step quantifies indirect effects either separately (each identified mediator) or simultaneously with a cumulated indirect effect of all mediators called overall indirect effect.

**Figure 1:**
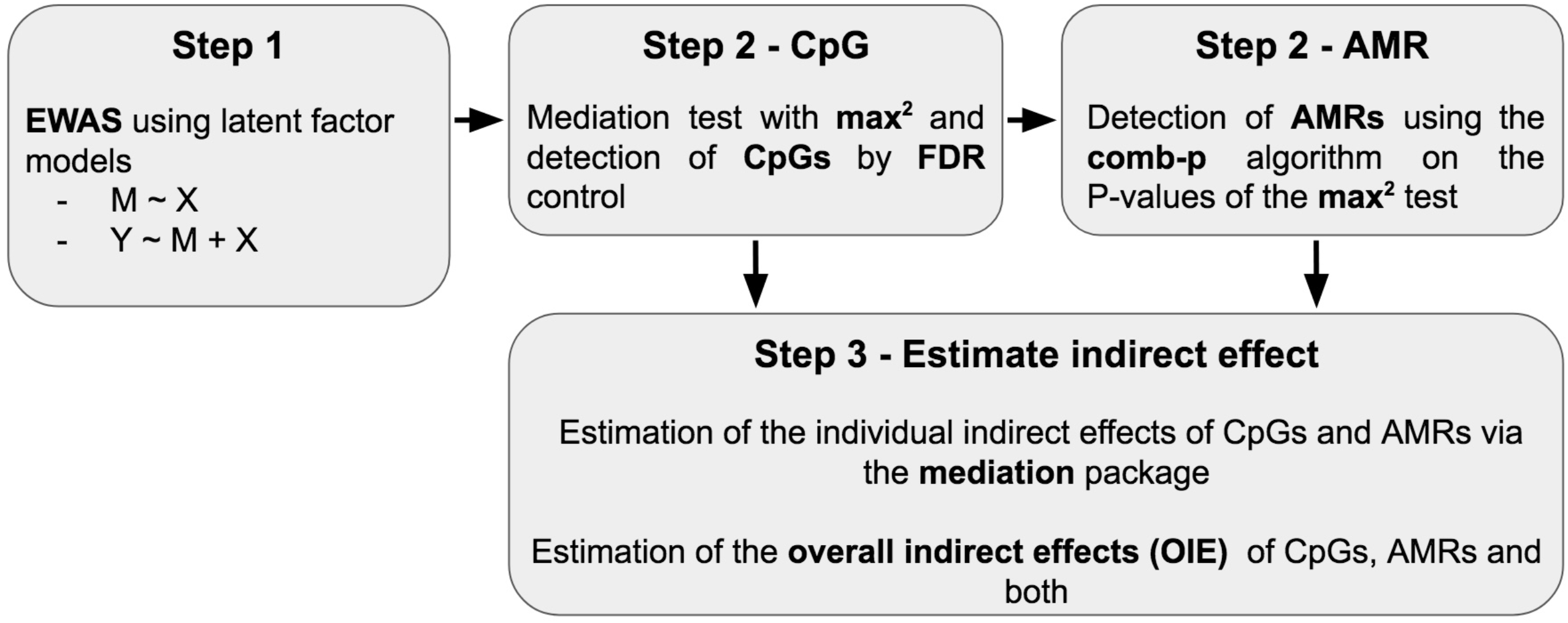
Workflow of the high dimensional mediation analysis procedure implemented in HDMAX2.

#### Step 1. Evaluating associations between exposure, mediators and outcome

The first step of HDMAX2 is to adjust latent factor mixed models (LFMMs) to estimate the effects of exposure, **X**, on a matrix **M** of CpG markers, and the effect of each marker on outcome, **Y**(24,25). LFMMs belong to a class of estimation algorithms that adjust latent factor models, and that encompass surrogate variable analysis (SVA) (26), directed SVA (27) or confounder adjusted testing and estimation (CATE) (28). Latent factor models differ from models based on a priori estimates of cell types (29,30), and represent a more general approach to the issue of confounding in association studies (26). Within the latent factor regression framework, additional known covariates, like maternal age or sex of the newborn, can be included in the model to improve accuracy.

Mediation analysis based on DNAm markers performs two EWAS. To estimate the effects of exposure (**X**) on a matrix of CpG markers (**M**), the following model was first adjusted to the centered data

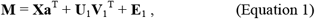

where **a** contains the vector of effect sizes of exposure on DNAm levels, **U**_1_ is a matrix formed of *K* latent factors estimated simultaneously with **a, V**_1_ contains the loadings associated with the latent factors, and **E**_1_ is a matrix of residual errors. The *K* latent factors represent hidden confounders, for example unobserved cell types of tissue samples and batch effects. Using the latent factor regression defined in Eq.1, a significance value, *P*x, is computed for the test of a null effect size for exposure on DNAm at each CpG marker (H0: *a*_j_ = 0, for the j^th^ marker).

A second EWAS was then performed in order to estimate effect sizes for the DNAm levels on the health outcome (**Y**) as follows:

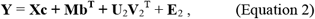

where **c** contains the direct effect of exposure on outcome, **b** contains the effect sizes of DNAm levels on outcome, **U**_2_ are latent factors from a latent factor regression model, **V**_2_ contains the corresponding loadings, and **E**_2_ is a matrix of errors. For each marker *j*, a significance value, *P*y, is computed for the test of a null effect size for DNAm on outcome (H0: *b*_*j*_ = 0, for the *j*^th^ marker).

#### Step 2. Identifying potential CpG mediators and aggregated mediator regions

The second step of HDMAX2 combines the significance values *P*x and *P*y computed at each DNAm marker by using a new procedure called the max-squared test. The *P*-value for the max-squared (max^2^) test was computed as *P* = max(*P*x, *P*y)^2^. Like the Sobel test, the max^2^ test rejects the null-hypothesis that either the effect of exposure on DNAm or the effect of DNAm on outcome is null. The square in the formula warrants that the distribution of *P*-values is uniform when *P*x and *P*y are independent and uniformly distributed. In HDMAX2, the max^2^ test was first used in order to identify potential CpG mediators. A combination of *P-* values along the methylome was then performed to identify potential AMRs using *comb-p*, a method relying on the Stouffer-Liptak-Kechris correction that combines adjacent CpG *P*-values in sliding windows (31). We considered methylated regions including at least two markers at a maximum distance of 1,000 bp and significant at the 10% False Discovery Rate (FDR) level. The mean value of DNAm levels for CpGs located in AMRs was retained to summarize information on methylated regions.

#### Step 3. Quantifying indirect effects with single and multiple mediators

Mediation of exposure on the outcome was first assessed at the level of CpG markers, and then at the level of aggregated regions. For CpG and for AMRs, estimates of indirect effect sizes and the proportion of mediated effect were computed in the R package mediation (8). For CpGs, the estimate of the indirect effect size for marker *j* was checked to be equivalent to the product of effect sizes,, computed in Eqs. (1) and (2). A novelty of HDMAX2 is to evaluate an overall (cumulated) indirect effect for all CpGs or AMRs identified in Step 3. The *overall indirect effect* (OIE) was estimated in a model including *m* mediator variables as follows

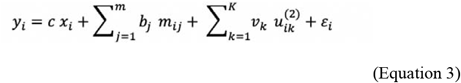

where () represent methylation levels observed at CpG mediators or AMRs (or both of them), and the terms () correspond to the latent factor coordinates estimated in Step 1. The overall indirect effect was then computed as

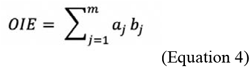

where (*a*_*j*_) represents the effect of exposure on methylation (Step 1). To account for correlation among mediators, the standard deviation of the OIE estimate was computed using a bootstrap approach (10,000 replicates).

### 2.2 Simulation studies

We performed simulations to compare the methods implemented in HDMAX2 with state-of-the-art approaches for EWAS in Step 1 and for mediation tests in Step 2.

#### Step-1 EWAS methods evaluated in simulations

In HDMAX2 Step 1, several latent factor estimation algorithms could be implemented for performing the EWAS. A preliminary study was performed to decide which of LFMM2, SVA and CATE was the best for our dataset using precision (1 - false discovery rate) and F1-score (harmonic mean of precision and power), as evaluation measures (Text S1, Figure S1). Then we performed generative simulations to compare methods using latent factors with those based on estimates of cell type composition. In this step, HDMAX2 was compared to two linear regression models including a priori estimates of cell type composition obtained from RefFreeEWAS (29) and refactor (30). Note that this a major difference with LFMM in which cell type composition is replaced by latent factor estimates computed simultaneously with effect size estimates (vectors **a** and **b** in Eqs. 1 and 2).

#### Step-2 mediation methods evaluated in simulations

We compared the max-squared mediation test in Step 2 of HDMAX2 to methods based on direct application of Sobel tests and of univariate mediation analysis (20,21). Then we compared the max-squared test to recent methods for high dimensional mediation: a multiple-testing procedure for high-dimensional mediation hypotheses, HDMT, similar to the max-squared test (10), a two-step familywise error rate procedure called ScreenMin (11), an approach using familywise error rate and false discovery rate control when testing multiple mediators SBMH (14), a linear regression model combined with an ANOVA (32) and an approach using variable selection to reduce the number of mediators HIMA (15). The last two approaches combined the first steps of HDMAX2 in a single step.

#### Mediation model simulations

The simulations were performed according to a generative model that reproduces the mediation pathways described in Eq. (1) and Eq. (2). Exposure and outcome (X and Y) and three confounding factors (**U**) were simulated according to a multivariate Gaussian model. The percentage of variance of exposure and outcome explained by the confounding factors, and the correlation between those variables, were set at 10%. The variances of confounding factors were equal to one. The number of DNAm markers was equal to *m* = 38,000, approximately equal to the number of CpGs for a single chromosome in our empirical data, and the number of individuals was equal to *n* = 500. The vectors of effect sizes (**a** for exposure and **b** for outcome) were generated by setting a proportion of effect sizes to zero. Non-null effect sizes were sampled according to a standard Gaussian distribution. A residual error matrix **E** was simulated by using a multivariate Gaussian distribution with means equal to zero and standard deviations of one. In addition to the three confounding factors, six additional factors representing distinct cell types were added in the simulation model. The proportion of cells from six different types were simulated by using a Dirichlet distribution. To consider values that are realistic with respect to our data analysis, the parameters of the Dirichlet distribution were equal to the proportions of each cell type estimated on the EDEN placental DNAm data (described afterwards). A matrix of DNAm markers was built using Eq 1 and Eq 2 with 3 parameters: the mean of non-null effect size for exposure (X) on methylation **M** (a = 0.2, 0.4), the mean of non-null effect size for **M** on outcome (b = 0.2, 0.4), and the number of putative causal markers (equal to 8, 16 or 32). For each set of parameters, 200 simulations were carried out. For each method tested, a subset of hits with a level of FDR = 5% was selected as potential mediators (33). For each list of hits, we computed precision (1 - FDR), sensitivity (power), and the harmonic mean of precision and sensitivity (F1-score). The highest value of an F1-score is one, if precision and sensitivity are maximal, and the lowest value is zero, if either the precision or the sensitivity is null.

### 2.3 Maternal smoking, placental DNA methylation, and pregnancy outcomes

#### Study population

Our analysis included participants of the EDEN Mother-Child Cohort enrolled in the university hospitals of Nancy and Poitiers, France, between 2003 and 2006 (34,35). Lifestyle, demographic, and clinical data were collected by questionnaires and interviews during pregnancy and after delivery.

#### DNAm measurements

DNAm was measured from DNA extracted from 668 placental samples. The Illumina’s Infinium HumanMethylation450 BeadChip was used to assess the levels of methylation in samples following the manufacturer’s instructions (Illumina, San Diego, CA, USA). Protocols for placental DNA extraction and DNAm processing are detailed in (36). Briefly, DNAm was normalized using the beta-mixture quantile (BMIQ) method to ultimately obtain beta-methylation levels for 379,904 CpG probed CpG sites (37).

#### Maternal smoking (MS), birth weight (BW) and gestational age (GA)

We excluded preterm deliveries (n=28), women who reported quitting smoking before pregnancy (n=70) and women whose smoking status was unknown (n=100), leaving 470 women included in our analyses. Birth weight was extracted from medical records. Prenatal maternal cigarette smoking was collected by questionnaires during prenatal and postpartum clinical examinations. *Non-smokers* were defined as women who did not smoke during the 3[months before and during pregnancy (359 non-smokers). *Smokers* were defined as women smoking more than one cigarette per day throughout the duration of the pregnancy (111 smokers). All smokers during pregnancy also smoked during the 3[months before pregnancy. *Gestational age* was defined as gestational age at birth.

#### Mediation analyses

We hypothesized that maternal smoking during pregnancy could induce modifications of placental DNAm that result in differences in GA or in BW. To this aim, we investigated the relationships between MS, placental DNAm and each pregnancy outcome. MS was encoded as a categorical variable and the outcomes were encoded as continuous variables. In order to identify mediators of the exposure-outcome relationship, we used the HDMAX2 approach to evaluate DNAm CpG mediators first, and then to identify AMRs.

In HDMAX2 regression models, adjustment factors included child sex, parity (0, 1, ≥□2 children; categorical covariate), maternal age at end of education (≤□18, 19–20, 21–22, 23– 24, ≥□25□years; categorical covariate), season of conception (categorical covariate), study center, maternal body mass index (BMI) before pregnancy, maternal age at delivery, batch, plate, and chip technical factors related to DNAm measurements (categorical covariates). We relied on the principal component analysis of the DNAm matrix to include 6 latent factors in the HDMAX2 regression models (Figure S2). This number was consistent with the 6 factors selected in a previous work to represent the cell-types using the Reffree algorithm (22). We adopted an empirical null approach, which can correct for shift in the data to respect the shape of the theoretical null (38). FDR-corrected *P*-values were calculated for the 379,904 CpGs using the local FDR algorithm in fdrtool (39). Calibration of the max^2^ test *P*-values was evaluated through a direct examination of the histogram of *P*-values. The local FDR parameter (eta0) was computed to evaluate the proportion of null-hypothesis among the 379,904 tests. This proportion was estimated at eta0 = 99.8-99.9%, suggesting that an FDR level of 5% would be overly conservative (Figure S3). To agree with the value of eta0, candidate CpGs were selected at FDR levels < 10%, corresponding to adjusted *P* < 9.03 × 10^−6^ for BW and to adjusted *P* < 3.27 × 10^−6^ for GA. Results obtained after considering FDR levels < 20% and < 5% are also reported.

#### Chained mediation of maternal smoking on birth weight

To better understand the causal pathways involving (six) genic regions that mediate the effect of MS both on GA and on BW, we hypothesized that GA has reverse effect on DNAm levels. To assess reverse causality, we evaluated the indirect effects of targeted AMRs in a mediation analysis of GA on BW and of BW on GA. For AMRs having a significant mediation *P*-value, each indirect effect and an overall indirect effect were computed from the above-described procedures.

#### Bioinformatic analyses

Promoter and enhancer regions were obtained from Illumina chip annotations. Gene annotations were obtained using the *FDb*.*InfiniumMethylation*.*hg19* package (40). Placental gene expression of annotated genes was compared to their gene expression in other tissues according to the Expression Atlas database (41). For every gene, Chauvenet’s criterion was used to decide whether the gene was outlier for placental expression compared to other tissues. Functional annotation was made from the KEGG and the Gene Annotation databases (42).

## 3. Results

### 3.1 Simulations

HDMAX2 was compared to several recent combinations of methods for multidimensional mediation analysis using simulation experiments. First, we compared the performances of latent factor models to other regression methods in estimating the association between exposure, DNAm levels, and outcome (Step 1 of HDMAX2). Then we compared the max^2^ mediation test to recently proposed tests (Step 2 of HDMAX2).

#### Performances of regression methods in step 1 of HDMAX2

A preliminary simulation study evaluated which of SVA, LFMM, or CATE provided the best estimation algorithm of latent factors for our empirical data set (Text S1). CATE and LFMM obtained better performance scores than SVA (Figure S1). LFMM runtimes were shorter than those of CATE, and LFMM performance scores were higher. Thus, we concluded that LFMM is the most appropriate for analysis of the EDEN cohort data, and we used it everywhere in subsequent assessments of HDMAX2. Using more general simulation experiments, we measured the relative performances of HDMAX2, that jointly estimates effect sizes and latent factors with LFMM, and linear regressions adjusted for a priori estimates of cell-type composition with RefFreeEWAS and ReFACTor (Figure 2). In all scenarios, the performances of the ReFACTor method were much lower than those of LFMM and RefFreeEWAS (Figure 2). For lower effect sizes of DNAm on outcome, LFMM and RefFreeEWAS reached close F1-scores, but LFMM obtained higher scores than RefFreeEWAS for higher effect sizes. All approaches obtained higher scores when more mediators were simulated, or when both the effect of exposure on DNAm and the effect of DNAm on outcome were higher. The results indicated that latent factor regression models outperformed methods that directly attempt to estimate cell-type composition from the DNAm data.

**Figure 2.**
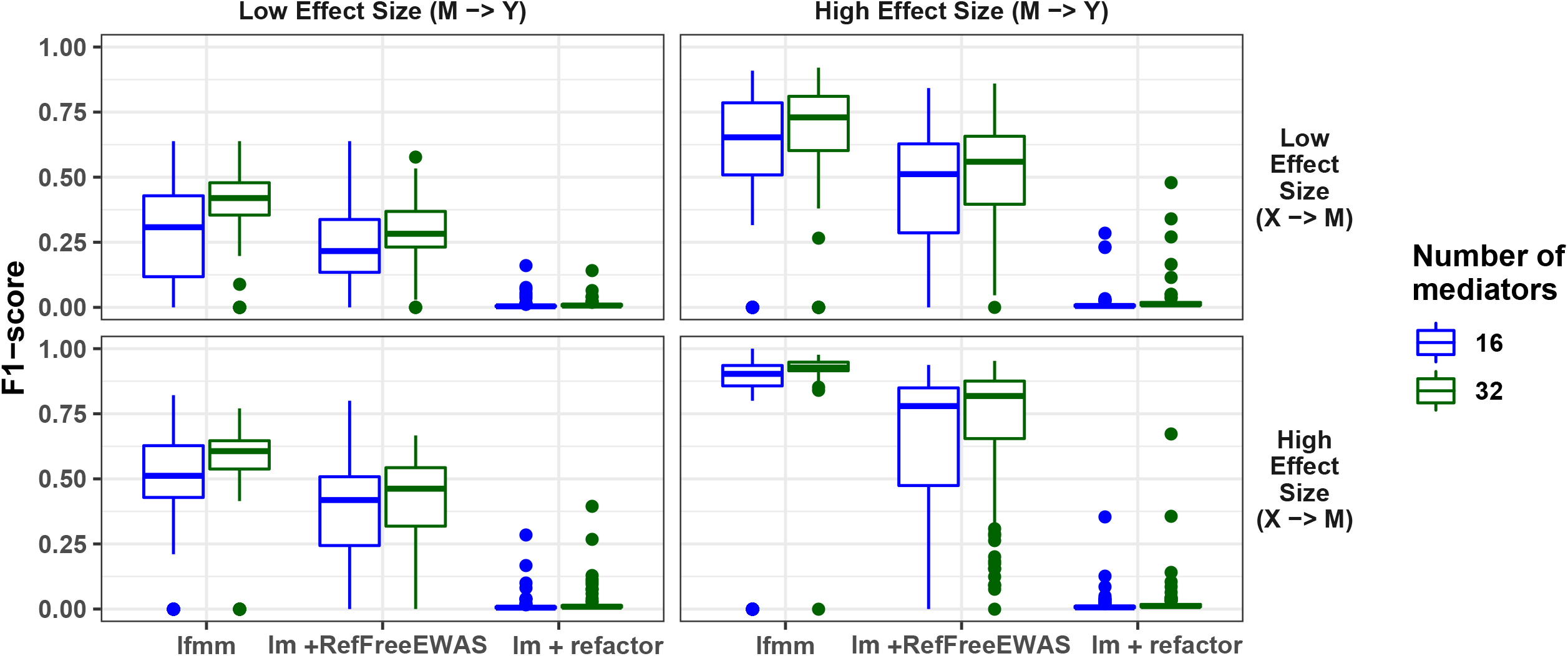
Relative performances of Step 1 approaches estimating latent factors versus inclusion of cell-type composition estimates. F1-score as a function of the number of mediators (16 or 32), effect size of exposure on DNAm (X->M; low = 0.2, high = 0.4), and effect size of DNAm on outcome (M->Y; low = 0.2, high = 0.4). Each simulation included 38,000 CpGs for 500 samples, with 6 cell types and 3 additional confounding factors.

#### Performances of mediation tests in the step 2 of HDMAX2

Next, we compared HDMAX2 to five recent tests for high-dimensional mediation: HDMT, ScreenMin, SBMH, linear models combined with ANOVA (lm+anova), and HIMA (Figure 3). In every scenario, HDMAX2 and HDMT reached similar scores, and those approaches were the best ones overall. In the specific case of high DNAm on outcome effect sizes and low exposure on DNAm effect sizes, lm+anova obtained the best scores, immediately followed by HDMAX2 and HDMT. The lowest performances were obtained with ScreenMin, SBMH and HIMA. When both effect sizes were high, HIMA obtained the lowest performances. For low DNAm on outcome effect sizes, lm+anova and SBMH obtained the poorest performances. In addition, HDMAX2 outperformed mediation analyses combining EWAS with Sobel tests and with unidimensional mediation analyses repeated at each marker, especially when the number of mediators increased from 16 to 32 (Figure S4). Since the runtime was much shorter for HDMAX2 than for HDMT and for other approaches (Figure S5), HDMAX2 was used in our analyses on empirical data.

**Figure 3:**
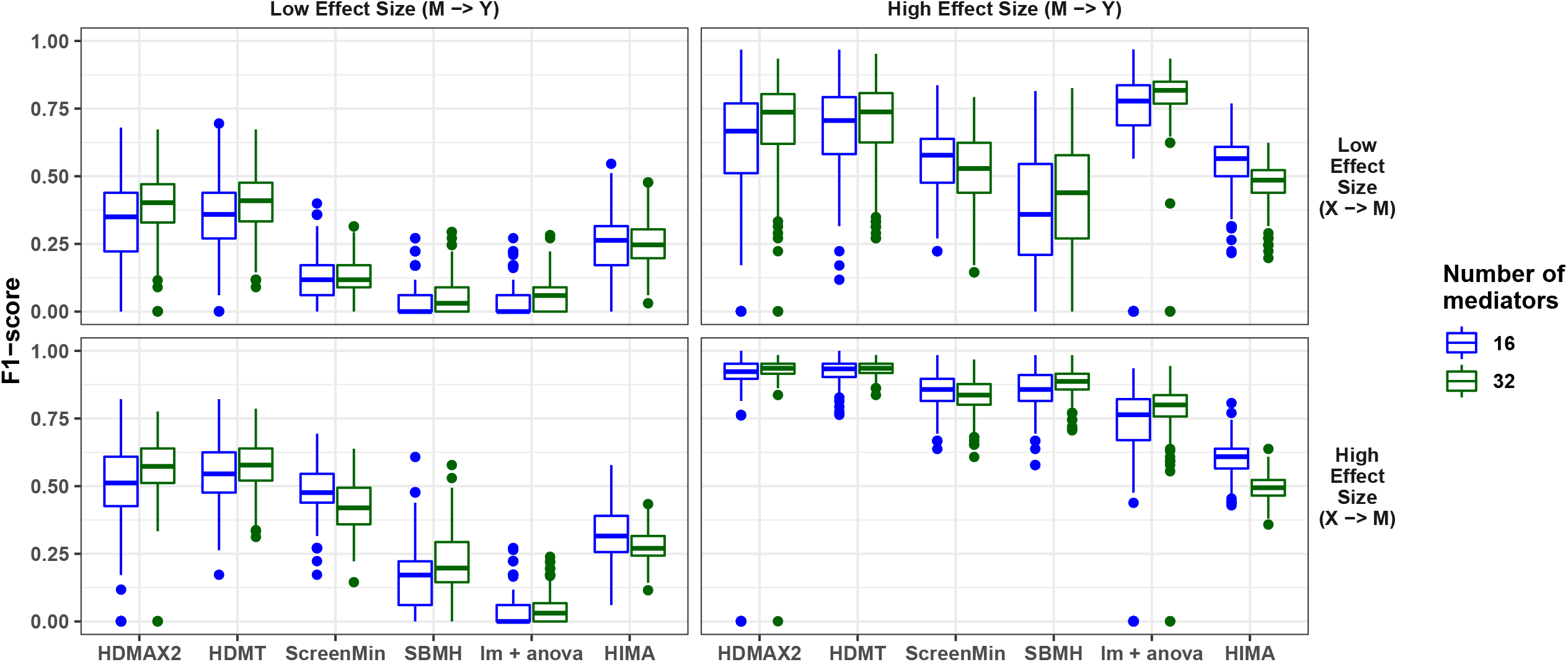
Relative performances of Step 2 multidimensional mediation methods. F1-score as a function of the number of mediators (16 or 32), effect size of exposure on DNAm (X->M; low = 0.2, high = 0.4), and effect size of DNAm on outcome (M->Y; low = 0.2, high = 0.4). Each simulation included 38,000 CpGs for 500 samples, with 6 cell types and 3 additional confounding factors.

### 3.2 Mediation of prenatal exposure to smoking on pregnancy outcomes

Among 470 mother-infant pairs, mean maternal age at enrolment was 29 years (SD = 5 years), body mass index before pregnancy was 23 kg/m^2^ (SD = 4.4 kg/m^2^) and 23.6% of women smoked during pregnancy (Table 1). Term birth weight (BW) ranged between 2010g and 4960 g, with a mean of 3352 g +/- 435 g. Gestational age (GA) varied from 37 weeks to 42 weeks, with a mean of 40 weeks +/-1.20 week (8.4 days). Maternal smoking (MS) during pregnancy had a significant correlation with BW (r = -0.16, *P* = 0.003), but not with GA (Figure S6). BW and GA were significantly correlated in mother-infant pairs (r = 0.31, *P* = 1.6 × 10^−12^). After adjustment, the total effect of MS was 140 g lower BW (SD = 49.1 g, *P* = 0.004), and the total effect of MS was not significant for GA (effect size = 0.12 week, SD = 0.14 week, *P* = 0.2434).

**Table 1.**
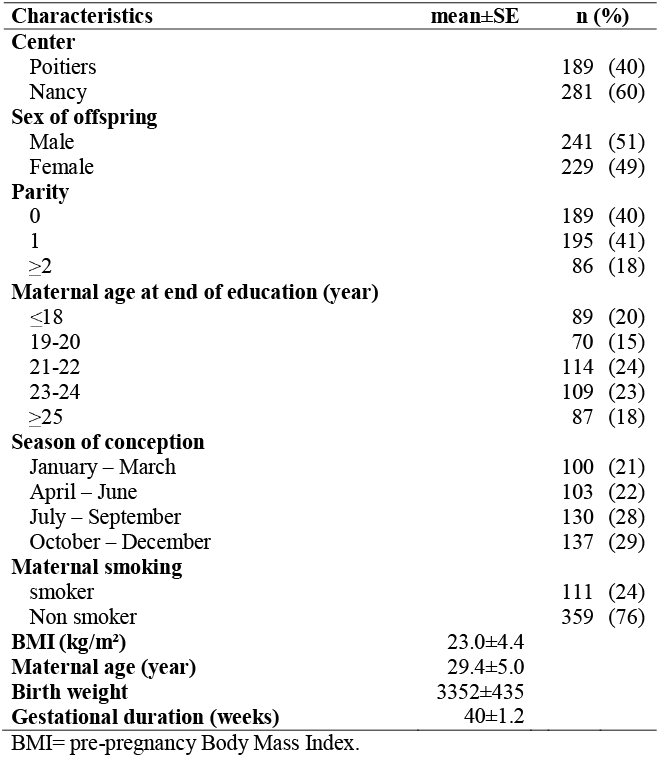
Characteristics of the study population (*n* = 470).

#### Mediation of maternal smoking on birth weight

A high dimensional mediation analysis of MS on BW was performed using placental DNAm data from the EDEN mother-child cohort. At an FDR level of 10% (5%), thirty-two (twenty) CpGs were identified as mediators of MS on BW (Table S1, Figure 4A, adjusted max-squared *P* < 9.11 × 10^−6^). Twenty CpGs were associated with a lower BW for the newborn (average ACME: -32.0 g, SD = 5.6 g; average proportion mediated (PM): 22.8%, SD = 4.0), and twelve CpGs were associated with a higher BW (average ACME: 32.6 g, SD = 10.3 g; average PM: 23.3 %, SD = 7.4) (Figure S7). The 32 CpGs were associated with an overall indirect effect corresponding to 40.3 g lower BW (SD = 51.3 g).

**Figure 4.**
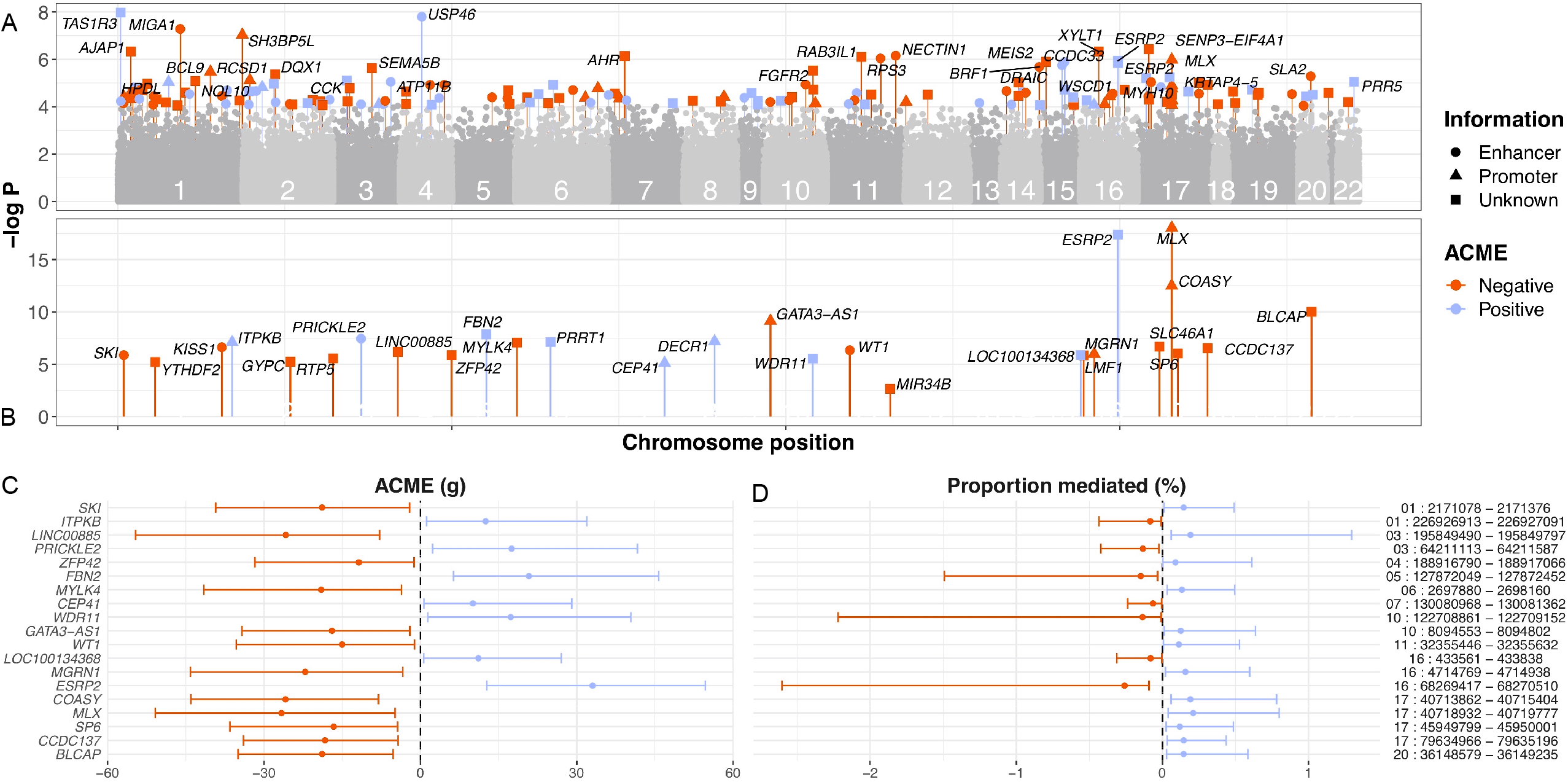
High Dimensional Mediation analysis (HDMAX2) of maternal smoking on birth weight. A) Manhattan plot for CpG’s -log(*P*-values) obtained from HDMAX2. Gene names correspond to hits identified at the 10% FDR level (32 hits). Colored bars without gene names correspond to hits identified at the 20% FDR level (164 hits). Gray bars without dots correspond to CpGs above the 20% FDR level. B) Manhattan plot of -log10(P-values) for potential AMRs at the 10% FDR level (28 hits). C) Estimates of indirect effect (ACME) and proportions of mediated effect for confirmed AMRs (19 hits). Symbols on top of colored bars correspond to classification as enhancer, promoter or unknown. Overall indirect effect of AMRs: 52 g lower BW.

Examples of CpG mediators with the largest negative indirect effects include cg10624729 (adjusted *P* = 5.15 × 10^−8^), in *MIGA1* (Mitoguardin 1) a regulator of mitochondrial fusion, associated with 41 g lower BW, cg19406975 (adjusted *P* = 9.27 × 10^−8^), in *SH3BP5L* (SH3 Binding Domain Protein 5 Like) which functions as a guanine exchange factor, associated with 41 g lower BW, cg01686933 (adjusted *P* = 6.98 × 10^−7^), in *NECTIN1* (Nectin Cell Adhesion Molecule 1) which encodes an adhesion protein that plays a role in the organization of epithelial and endothelial cells, associated with 41 g lower BW, and cg14502606 (adjusted *P* = 1.04 × 10^−6^), in *MLX* (MAX Dimerization Protein MLX) a transcription factor which plays a role in proliferation, determination and differentiation, associated with 38 g lower BW (Table S1).

At an FDR level < 20%, 164 mediators were discovered, including fifty-five CpGs within enhancer regions and twenty-six CpGs within promoter regions (Figure 4A). In comparison with the methylome, the list of mediators was enriched in hits corresponding to enhancer regions (33% of all hits, *P* = 0.0003, Fisher test, Figure S8A), and it was depleted in hits corresponding to promoter regions (15% of all hits, *P* = 0.04, Fisher test, Figure S8B). Several mediators were found in the body of a gene (109 hits), and some genes were hit more than once (*AJAP1, ESRP2, SH3BP2, SKI, SRSF5, VAV2* and *MLX*). We additionally performed mediation analyses for CpG cg27402634 (between *LINC00086* and *LEKR1*) and cg25585967 (*TRIO*) identified in (Morales et al. 2016), and for one CpG (cg11280108) in the HumanMethylation450 BeadChip which was among the seven CpGs identified in (Cardenas et al. 2019) from the EPIC chip. Although associations of DNAm with exposure to MS were significant for those CpGs (adjusted *P* = 9.07 × 10^−14^), none of those markers were mediators of MS on BW in our analysis (*q-*values > 0.93).

Regarding methylated regions, HDMAX2 detected twenty-eight potential AMRs, including four within enhancer regions, seven within promoter regions, and twenty within the body of a gene (FDR level < 10%, Figure 4B). Nineteen AMRs were associated with statistically significant indirect effects ranging between 26.7 g lower BW and 33.0 g higher BW (Table S2). Twelve AMRs were associated with a lower BW (average ACME: -19.7 g, SD = 4.6; average PM: 14.0%, SD = 3.3%), and seven were associated with a higher BW (average ACME: 17.5 g, SD = 7.9; average PM: 12.5%, SD = 5.6%, Figure 4C). The 19 AMRs were associated with an overall indirect effect corresponding to 52 g lower BW (SD = 45 g). The overall indirect effect of both CpG mediators and AMRs was 44.5 g lower BW (SD = 60.7 g). The strongest evidence corresponded to AMR chr17:40,713,862-40,715,404 (adjusted *P* = 3.20 × 10^−13^) in *COASY* (Coenzyme A Synthase) which plays an important role in numerous synthetic and degradative metabolic pathways in all organisms, associated with 26 g lower BW. This AMR was only 3kb close to another AMR, chr17:40,718,932-40,719,777 (adjusted *P* = 9.37 × 10^−19^), in *MLX*, which was associated with 27 g lower BW (Figure 4, Table S2 for a full list of AMRs).

#### Mediation of maternal smoking on gestational age

An independent mediation analysis was performed on the DNAm data in order to evaluate the indirect effects of MS on GA. At an FDR level < 10%, fifteen CpGs (two CpGs at FDR level < 5%) were identified as mediators of MS on GA (Table S3, Figure 5A, adjusted max-squared *P* < 3.28 × 10^−6^). The 15 CpGs were associated with a weak overall indirect effect corresponding to 0.28 week (2 days) lower GA (SD = 0.12) (Figure S9).

**Figure 5.**
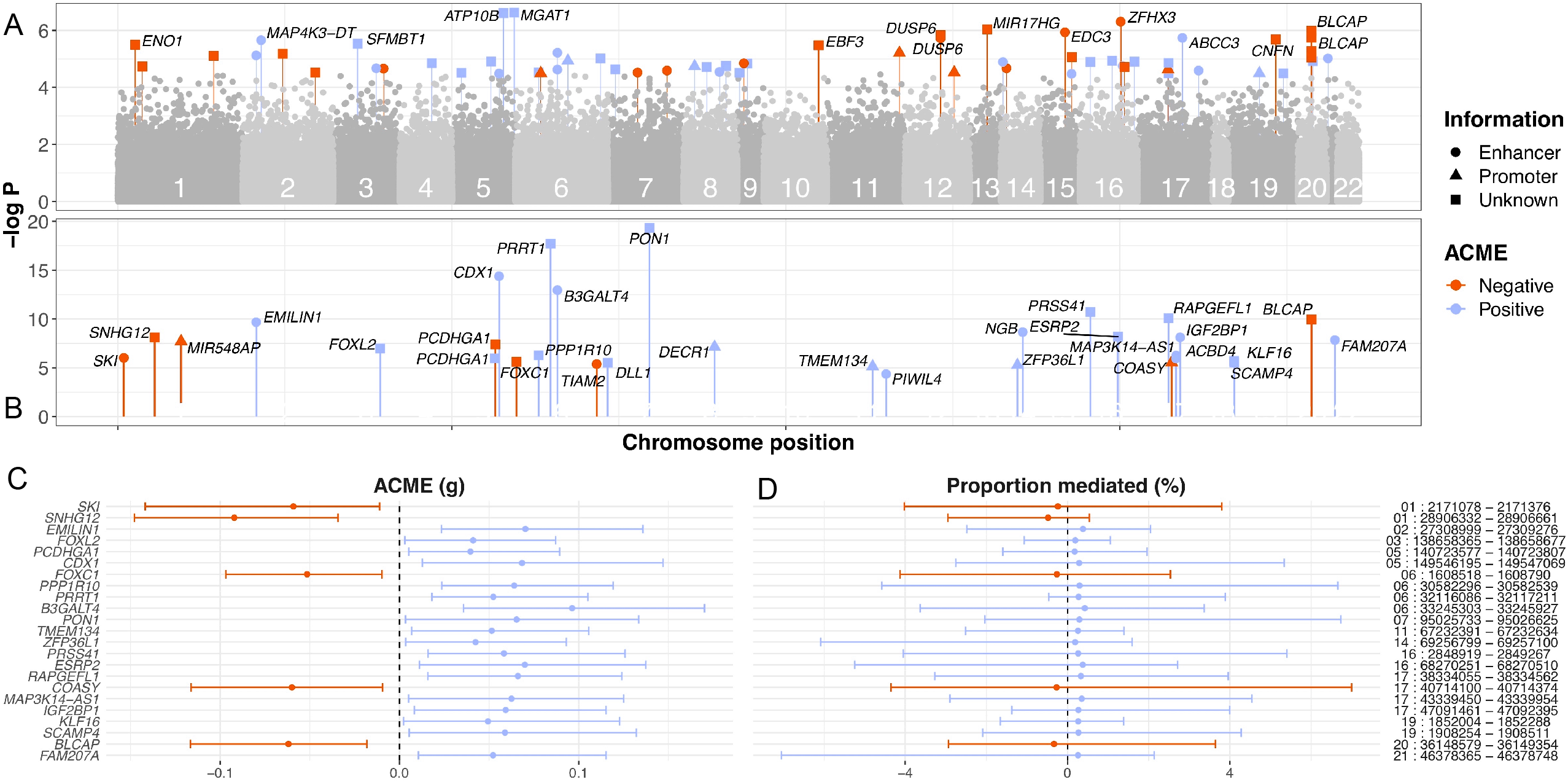
High dimensional Mediation analysis (HDMAX2) of maternal smoking on gestational age. A) Manhattan plot for CpG’s -log(*P*-values) obtained from HDMAX2. Gene names correspond to hits identified at the 10% FDR level (15 hits). Colored bars without gene names correspond to hits identified at the 20% FDR level (63 hits). Gray bars without dots correspond to CpGs above the 20% FDR level. B) Manhattan plot of -log10(P-values) for potential AMRs at 10% FDR level (31 hits). C) Estimates of indirect effects (ACME) and proportions of mediated effect for confirmed AMRs (23 hits). Symbols on top of colored bars correspond to classification as enhancer, promoter, or unknown. Overall indirect effect of AMRs: 0.12 weeks lower GA.

Examples of CpG mediators with the most negative effects include cg10298741 (adjusted *P* = 4.82 × 10^−7^), in *ZFHX3* (Zinc Finger Homeobox 3) a transcription factor which regulates myogenic and neuronal differentiation, associated with 0.08 week lower GA, cg04908961 (adjusted *P* = 9.19 × 10^−7^), in *MIR17HG* (MiR-17-92a-1 Cluster Host Gene) a host gene for the MiR17-92 cluster, a group microRNAs (miRNAs) that may be involved in cell survival, proliferation, and differentiation, associated with 0.09 week lower GA, cg08402058 (adjusted *P* = 1.04 × 10^−6^), in *BLCAP* (Bladder cancer-associated protein) which reduces cell growth by stimulating apoptosis, associated with 0.09 week lower GA, (see Table S3 for a full list of CpG mediators). Ten CpGs were associated with a shorter GA (average indirect effect 0.09 week lower GA, SD=0.02; PM: 74%, SD = 14%), and five CpGs were associated with higher GA (average ACME: 0.09 week, SD = 0.01; PM: 71%, SD = 10%; Figure S9). At an FDR level < 20%, sixty-three mediators were identified, including twenty-six hits within an enhancer region (Figure 5A). This subset of CpG mediators was enriched in hits corresponding to enhancer regions (33% of all hits, P < 2.2 × 10^−16^, Fisher test, Figure S8A).

The per-region analysis resulted in the detection of thirty-one potential AMRs, including eleven regions within enhancers, five within promoters, and twenty-six within the body of a gene (Figure 5B, Table S4). Twenty-three AMRs were associated with small but statistically significant indirect effects ranging between -0.09 week and 0.10 week (none were associated with a significant mediated proportion). Five regions were associated with a lower GA (average ACME: -0.06 week ; SD = 0.01 ; average PM: 54.1%, SD = 13.1%), and eighteen regions were associated with a higher GA (average ACME : 0.06 week ; SD = 0.01; average PM: 49.4%, SD = 11.1%).

The 23 AMRs were associated with a weak overall indirect effect corresponding to 0.12 week (23 hours) shorter GA (SD = 0.11). The cumulative overall indirect effect of CpG mediators and AMRs was 0.09 week (16 hours) shorter GA (SD = 0.14). The largest negative indirect effects corresponded to AMR chr1:28,906,332 - 28,906,661 (adjusted *P* = 7.89 × 10^−9^) in *SNHG12*, (Small Nucleolar RNA Host Gene 12) an RNA gene that may promote tumorigenesis, associated with 0.09 week lower GA, chr20:36,148,579 - 36,149,354 (adjusted *P* = 1.13 × 10^−10^) in *BLCAP*, which encodes a protein that reduces cell growth by stimulating apoptosis, associated with 0.06 week lower GA, and chr17:40,714,100 - 40,714,374 (adjusted *P* = 2.84 × 10^−6^) in *COASY* associated with 0.06 week lower (Figure 5, Table S4 for a full list of AMRs).

#### Chained mediation of maternal smoking on birth weight through DNAm and GA

Six genes, *COASY, BLCAP, SKI, DECR1, ESRP2, PRRT1*, included AMRs that act as mediators both for MS on BW and for MS on GA. To better understand the causal pathways involving those genic regions, we tested the hypothesis that GA influences methylation levels in those regions, and estimated the indirect effects in a mediation analysis of GA on BW (Figure 6, Figure S10, Table S5). In this analysis, GA had significant indirect effects on BW for two of the six AMRs, in *COASY* (ACME = 6.9 g, mediation *P* <10^−3^), *BLCAP* (ACME = 5.1 g, mediation *P* = 0.01). The two AMRs were associated with an overall indirect effect corresponding to 10 g higher BW (SD = 3.91). We found no evidence that one of these 6 AMRs was present in the pathway from BW to GA.

**Figure 6.**
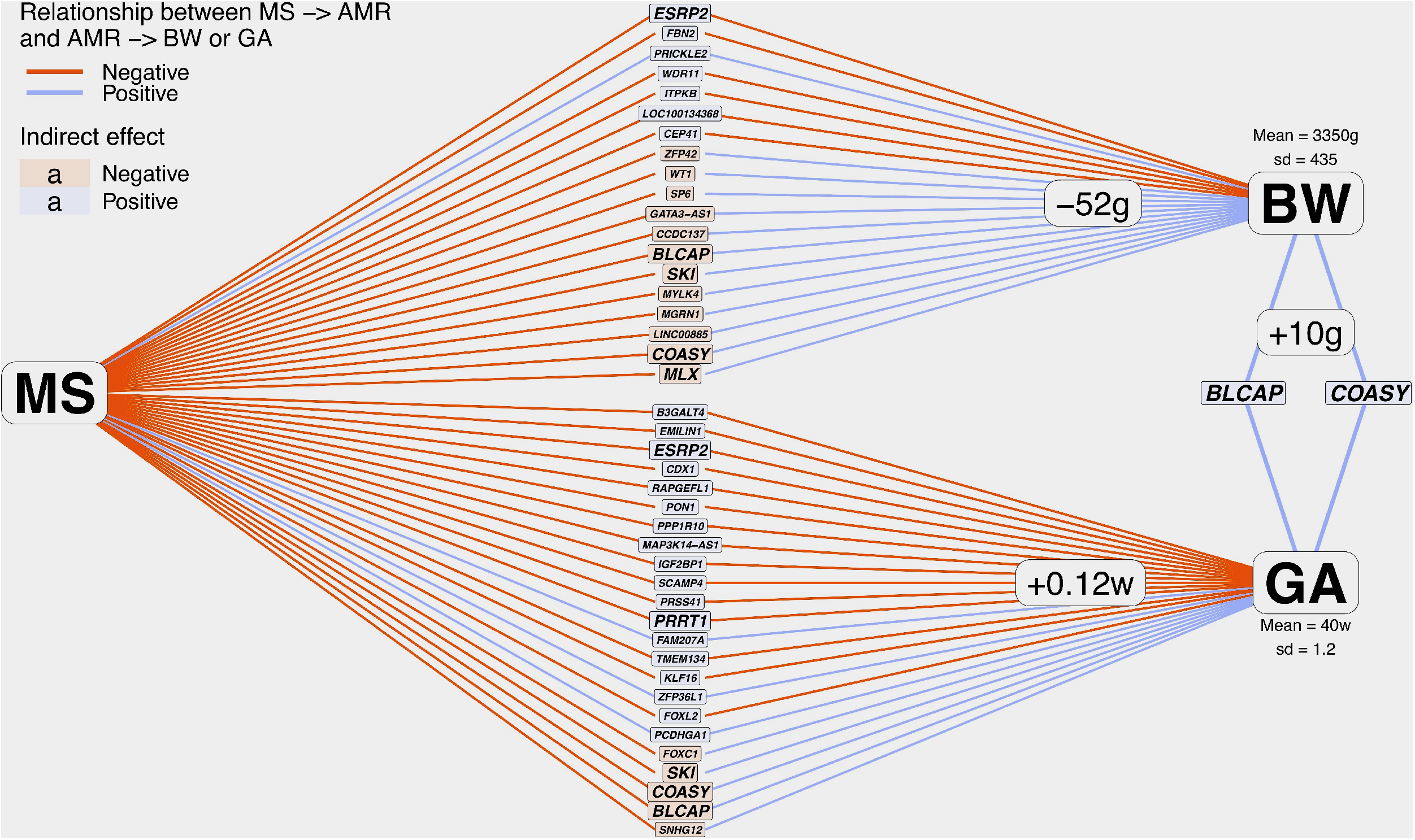
Summary of mediation analysis of MS on GA and BW for AMRs. Nineteen AMRs mediate the relationship between MS and BW, with a total indirect effect corresponding to 52 g lower BW. Twenty-three AMRs mediate the relationship between MS and GA, with a total indirect effect corresponding to 0.12 weeks shorter GA. Two AMRs mediate the relationship between GA and BW with a total indirect effect corresponding to 10 g higher BW. The color of each gene’s box indicates the sign of the indirect effect. The color of segments indicates the direction of association (blue means positive, orange means negative).

In the genomic region surrounding the *COASY* gene (Figure S10), the AMRs were located in regions with low DNAm levels (Figure S11B), and MS decreased DNAm levels within AMRs (Figure S11C). The CpGs contained in AMRs mediated lower BW, and were among the most negative observed indirect effects (Figure S11D-E). In the genomic region surrounding the *BLCAP* gene (Figure S12), AMRs were located in highly methylated gene body areas (Figure S12B), and MS decreased DNAm levels within AMRs (Figure S12C). The CpGs contained in AMRs mediated lower BW, and again, they were among the most negative observed indirect effects (Figure S12D-E). Figure 6 provides a summary of the chained mediation analysis (Figure S13 for a summary of CpG mediation analysis).

## 4. Discussion

### Main contributions

High dimensional mediation analysis holds promising results for deciphering molecular mechanisms underlying the association between exposure and outcomes. We presented HDMAX2, a method combining estimates of latent factors in EWAS with max-squared tests for mediation, which also evaluates an overall mediated effect for CpG or AMR. Using simulations, we performed an in-depth evaluation of the statistical performances of HDMAX2, and showed that HDMAX2 outperforms state-of-art methods and recent approaches proposed to identify mediators in a high dimensional setting. HDMAX2 was applied to assess the indirect effects of exposure to MS on GA and BW in a study of 470 women from the EDEN mother-child cohort, and confirmed the important role played by placental DNAm in the pathway between maternal smoking during pregnancy and fetal growth outcomes (3). In addition to single CpG mediators, our analysis examined AMRs and computed an overall indirect effect of all mediators considered simultaneously. The overall indirect effects of CpG and AMR were 44.5 g lower BW (32.1% of the total effect size) and 0.09 week lower GA (75% of the total effect size). These results support the hypothesis that the role of placental DNAm in the mediation of effect of exposure to MS on BW and on GA may be more polygenic than previously reported. In addition, a chained mediation analysis of MS on BW suggested the existence of reverse causal relationships for AMR located in the genes *COASY* and *BLCAP*, which mediate a proportion of the effect of MS on BW through an effect of GA on DNAm.

### Simulation studies

The main improvements of HDMAX2 over existing mediation methods is the use of latent factor models for estimating hidden confounders in step 1, and the max^2^ test of mediation in step 2. The combination of latent factors and max^2^ tests proposed by the HDMAX2 approach was carefully evaluated with intensive simulations, and resulted in increased performances compared to five state-of-the-art methods evaluating multiple mediators (10,11,14,15,32). Latent factors increased statistical power compared to using a priori estimates of cell-type proportions from reference-free methods (29,30). The max-squared tests showed considerably better performances in comparison to the univariate mediation or Sobel test approaches, which were used in previous studies analyzing the role of placental DNAm data in the pathway between MS and BW (20,21). Using HDMAX2, none of the mediating CpGs identified using univariate mediation or Sobel test approaches (Morales et al. 2016 Cardenas et al. 2019) were mediators of MS on BW in our analysis (*q-* values > 0.93).

### Mediation analysis of maternal smoking on birth weight

Previous studies have shown a possibly overestimated mediated effect of MS on BW, sometimes greater than the total effect size (43). This is a limitation of univariate indirect effects estimated independently in a context of correlation between multiple mediators. In contrast, our approach estimated an overall indirect effect of the placental methylome representing 32% of the total effect size of MS on BW. Compared with previous placental DNAm mediation analyses of MS on BW (Cardenas et al., 2019; Morales et al., 2016), the magnitude of each mediator indirect effect size estimated in our cohort represented smaller part (less than 24% for AMRs) of the total effect size, and it was spread over more mediators suggesting that indirect effects are more polygenic than in previous estimates.

### CpG mediators

HDMAX2 identified 32 CpG mediators of MS on BW, for which a majority (20/32) of effects represented a lower BW. The results provided evidence for an enrichment in enhancer regions and for a depletion in promoter regions among mediators, which agrees with conclusions from an association study between MS and placental DNAm in the EDEN cohort (22). According to the Gene Ontology database, six mediators were located in genes linked to development or to the growth of tissues: cg24571086 in *FGFR2*, cg11362604 in *MEIS2*, cg00108098 in *SEMA5B*, cg10778780 in *CCK*, and cg20482145 in *MYH10* and cg07156115 in *AHR*. The genes *FGFR2* and *SEMA5B* are linked to the development of multicellular organisms and to the growth of developmental organs, *MEIS2* is linked to the development of the brain, eyes and pancreas, *CCK* is linked to neuron migration, and *AHR* is linked to the development of blood vessels.

### Aggregated mediator regions

Evidence for increased polygenicity of placental DNAm mediation was confirmed by examination of AMRs, which are seen as more robust and more biologically meaningful than isolated differentially methylated CpGs (44). HDMAX2 identified 19 AMRs of MS on BW, for which a majority of effects represented a lower BW (Figure 4C, Table S2). The most negative effects corresponded to AMRs in *COASY*, which plays an important role in numerous synthetic and degradative metabolic pathways and in *MLX*, a transcription factor physically close to *COASY*, which is co-expressed in the placenta.

Four regions were located in genes linked to tissue development or growth, in *FBN2* related to camera-type eye development, *ZFP42* to gonad development, *ESRP2* to fibroblast growth factor receptor signaling pathway, and *SKI* to roof of mouth development, olfactory bulb development, camera-type eye development, and skeletal muscle fibber development. The genes *FBN2* and *ZFP42* were over-expressed in the placenta compared to other tissues. Smoking-induced AMRs in *FBN2* and *ESRP2* were associated with higher BW, whereas AMRs in *ZFP42* and *SKI* were associated with lower BW. Looking more closely at the biology of mediators, we found a large number of them located in genes related to preeclampsia, a pregnancy complication of placental origin characterized by high blood pressure and protein in the urine, causing about a third of very premature births. Preeclampsia-related genes included *NECTIN1* (45), *AHR* (46), *FGFR2* (47), *COASY* (48), *BLCAP* (49), *SKI* (48), *AJAP1* (50), *SH3BP5* (51). The over-representation of preeclampsia-related genes supports a pleiotropic effect of mediators, and highlights the difficulty of disentangling relationships between correlated outcomes.

### Mediation analysis of maternal smoking on gestational age and potential for reverse causality

Our results provided evidence that DNAm (CpG + AMR) mediates a relatively small total indirect effect of MS on GA, representing 0.09 week lower GA (15 hours). The largest negative effects corresponded to AMRs located in *SNHG12* and in *BLCAP* (Table S4). Six genes contained DMRs mediating both the effect of MS on BW and the effect of MS on GA. Two of those AMRs, located in *BLCAP* and *COASY*, had among the largest negative effects on both GA and BW. We reported strong evidence that *BLCAP* and *COASY* were present in the pathway from GA to BW, but no evidence that they were present in the pathway from BW to GA. This result indicates that the corresponding AMRs in *BLCAP* and *COASY* may be involved in complex causal relationships, in which DNAm plays a role in the negative effect of MS on BW (and on GA), which is amplified by a lower GA (Figure 6). Knowing whether placental DNAm influences GA or GA influences placental DNAm remains an open question. A limitation to interpretation is the fact that GA and placental DNAm are co-occurring events. However, our results suggest a bidirectional association between placental DNAm and GA, with a feedback loop from GA to BW through placental DNAm.

### Universally applicable framework for high dimensional mediating events

A large body of epigenetic research in perinatal health is dedicated to cord blood DNA methylation, although the placenta has attracted recent attention (20,21,52). The placenta exhibits a unique epigenetic profile, as it is one of the tissues with lower DNA methylation levels, which undergoes intense remodeling in early gestation, and dynamic changes with increased DNA methylation as gestation advances (53,54).The placenta supports both the health of the mother and the development of the foetus: it produces hormones, ensures immune-tolerance, provides nutrients to the foetus and regulates the exchange of gases and wastes. The placenta contains key information on the intra uterine environment, and is a highly relevant tissue to investigate within the DOHAD framework. Besides being associated with several prenatal exposures, placental DNA methylation is suggested to be a relevant proxy for neurodevelopmental outcomes (55–57) and respiratory health (58) of the child. Understanding the indirect effects of placental DNAm modifications on such outcomes will be an important objective, for which the HDMAX2 framework will be very helpful. Beyond the role of the placenta and DNA methylation, other tissues and omics markers are relevant to investigate in perinatal and more generally epidemiological studies. The HDMAX2 framework can be applied with other layer of mediators, basically any type of high-throughput data (i.e. gene expression data), or with data on any other tissue types.

### Summary

We developed a novel algorithm for high-dimensional mediation, HDMAX2. Beyond our current application to placental DNAm data, HDMAX2 is applicable to a wide range of tissues and omic layers including genomics, transcriptomics, and other types of omics. HDMAX2 showed better performances on simulations and increased power compared to existing approaches. We showed the strength of HDMAX2 by applying it to characterize associations between exposure to maternal smoking during pregnancy and birth weight and gestational age at birth of the baby. The mediation analysis suggested a causal relationship between maternal smoking during pregnancy and those outcomes underpinning many more epigenetic regions than previously found, suggesting a polygenic architecture for the pathways. Not limited to single CpG markers, HDMAX2 is extended to identifying AMRs. AMRs provide more robust evidence than single CpGs, and allowed the characterization of regions mediating effects of maternal smoking during pregnancy both on gestational age and birth weight, suggesting that placental DNAm is an important biological mechanism. We further showed the overall indirect effect accounting simultaneously for all mediators identified as a plausible estimate of the mediated effect. AMRs located in *COASY* and *BLCAP* suggested reverse causality in the relationship between gestational and the methylome contributing to lower birth weight. Our study added several statistical improvements to high-dimensional mediation analyses, and revealed an unsuspected complexity of the causal relationships between maternal smoking during pregnancy and birth weight at the epigenome-wide level. Limitations of the current work, and thus future research avenues, include a better characterization of interactions and of the polygenic architecture of phenotypes, especially when there is a high number of markers with small-effect-sizes, which will require much larger sample sizes (59).

## Data Availability

The EDEN datasets analyzed in the presented study are not publicly available as they are containing information that could compromise the research participants privacy/consent. However, they are available from the corresponding author on reasonable request and with permission from the EDEN Steering Committee.

## Acknowledgments

We thank Daniel Vaiman (INSERM U1016) for his help with lab experiments. We thank all the participants and members of the EDEN mother-child cohort study group.

## Abbreviations

ACME: Average causal mediation effects
AMR: Aggregated Mediator Region
BMI: Body Mass Index
BMIQ: Beta Mixture Quantile
BW: Birth Weight of the baby
CATE: Confounder adjusted testing and estimation
CpG: Cytosine-phosphate-Guanine
DNAm: DNA methylation
DOHaD: Development Origins of Health and Diseases
EWAS: Epigenome Wide Association Studies
FDR: False Discovery Rate
GA: Gestational Age at delivery
HDMAX2: High-Dimensional Mediation Analysis with max-squared tests
HDMT: High-Dimensional Mediation Test
LFMM: Latent Factor Mixed Model
MS: Maternal Smoking during pregnancy
OIE: overall indirect effect
PM: Proportion Mediated
SVA: Surrogate Variable Analysis

## 5. Supplementary information

### See supplemental materials including tables and figures

#### Software availability and requirements

The method presented in this study is available in the R package HDMAX2 at https://github.com/bcm-uga/hdmax2

GNU and reusable under General Public License v3.0. Scripts reproducing the simulations analyses are available at https://github.com/bcm-uga/HDMAX2_Simulation_Scripts. The R package lfmm is publicly available from CRAN.

### Funding

This work was supported by a grant from the French National Cancer Institute (INCa), the French Institute for Public Health Research (IreSP) (INCa_13641), and the French Agency for National Research (*ETAPE*, ANR-18-CE36-0005). BJ was partly supported by the Grenoble Alpes Data Institute, supported by the French National Research Agency under the Investissements d’Avenir program (ANR-15-IDEX-02), and by LabEx PERSYVAL Lab, ANR-11-LABX-0025-01. DNA methylation measurements were obtained thanks to grants from the Fondation de France (no. 2012-00031593 and 2012-00031617) and the French Agency for National Research (ANR-13-CESA-0011).

The EDEN mother-child study was supported by Foundation for medical research (FRM), National Agency for Research (ANR), National Institute for Research in Public health (IRESP), French Ministry of Health (DGS), French Ministry of Research, INSERM Bone and Joint Diseases National Research (PRO-A), and Human Nutrition National Research Programs, Nestlé, French National Institute for Population Health Surveillance (InVS), French National Institute for Health Education (INPES), the European Union FP7 programmes (FP7/2007-2013, HELIX, ESCAPE, ENRIECO, Medall projects), Diabetes National Research Program, French Agency for Environmental Health Safety (ANSES), Mutuelle Générale de l’Education Nationale (MGEN), French national agency for food security, Frenchspeaking association for the study of diabetes and metabolism (ALFEDIAM).

### Author’s contributions

BJ, CCB, ME performed the statistical analyses. JL, OF designed the study, wrote the manuscript and obtained funding. OF developed the statistical analysis plan. BH supervised the data collection and data management of the EDEN cohort, and provided guidance on the project. JT supervised the methylation and lab arrays. All authors read, revised and approved the final manuscript.

### Availability of data and materials

The EDEN datasets analyzed in the presented study are not publicly available as they are containing information that could compromise the research participant’s privacy/consent. However, they are available from the corresponding author on reasonable request and with permission from the EDEN Steering Committee.

### Ethics approval and consent to participate

The EDEN cohort received approval from the ethics committee (CCPPRB) of Kremlin Bicêtre and from the French data privacy institution Commission Nationale de l’Informatique et des Libertés (CNIL). Written consent was obtained from the mother for herself and for the offspring.

### Consent for publication

Not applicable.

