## Supplementary material for "High Dimensional Mediation Analysis: a new method applied to maternal smoking, placental DNA methylation and birth outcomes": Supplmental material

### **Supplementary materials**

Text S1. **Simulation study of EWAS based on latent factor regression models and the EDEN cohort data set.**

We used a conditional simulation algorithm to simulate data used in EWA studies and to evaluate the performance of statistical methods for latent factor regression models as defined in equation (1) of the main text. We implemented a simple linear model (t-test), and three other estimation algorithms: surrogate variable analysis (SVA, Leek et al. 2012), confounder adjusted testing and estimation (CATE, Wang et al. 2017) and latent factor mixed models 2 (LFMM2, Caye et al. 2019), and relied on the performances of these methods on our simulated data set to implement the best performing algorithm in the HDMA approach.

Consider a matrix of methylation profiles, ***M***, obtained for *n* individuals and corresponding to *M* markers. We assume that regression models, *m_j_* = *b_j_X* + *E_j_*, describes the relationship between an unobserved exposure variable, *X,* and methylation levels observed for the *j*-th marker (*b_j_* is the size of the effect on exposure on methylation level *j*, and *E_j_* has variance *σ_j_*^2^). Conditional on the matrix of methylation profiles, we can simulate the unobserved exposure *X*, of variance *σ_X_*^2^, as follows

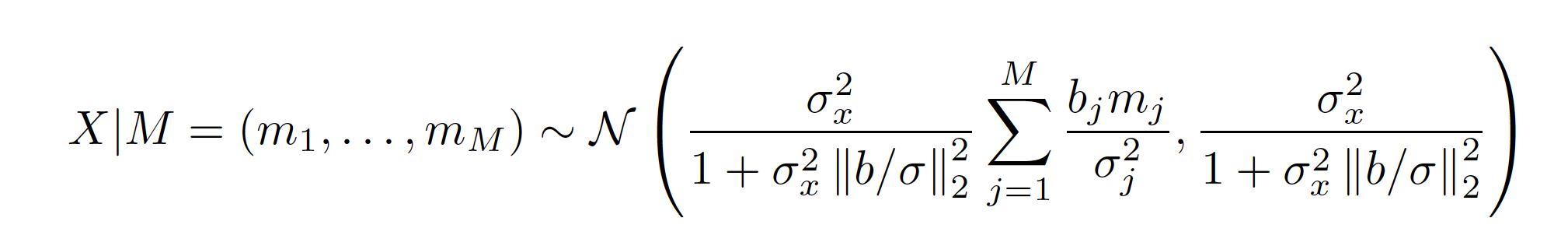

where

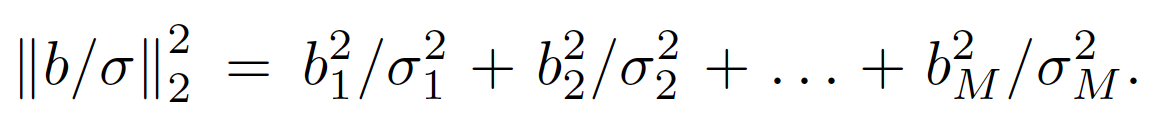

Using data from chromosome 1 in the EDEN cohort data of placenta DNA methylation (see main text 2.3), we defined causal markers, for which *b_j_* is non-null, and the corresponding effects on methylation levels as follows. One hundred EWAS were generated from those data with 30 randomly chosen causal markers and *σ_x_* = 0*.*18. The effect sizes at causal markers were defined as *b_j_ = σ_j_* /√0.1. In agreement with the data analysis performed in the main text, we chose k = 5 latent factors in SVA, CATE and LFMM2. Statistical performances were evaluated by the precision (one minus the false discovery rate) and F-score (harmonic average of precision and power). The results provide evidence that LFMM2 and CATE performed better than SVA and the simple linear model (Figure S1).

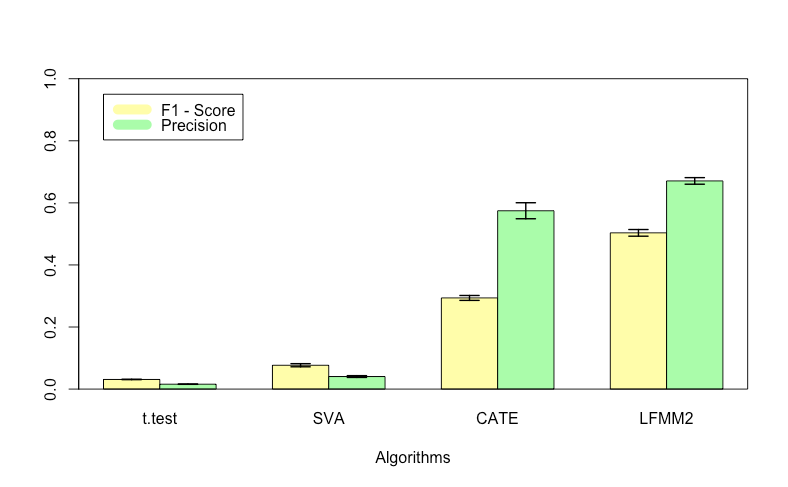
**Figure S1. Statistical performances of four regression models in simulated EWAS.** Exposure data were simulated conditional on observed data sampled from the EDEN cohort. SVA: Surrogate Variable Analysis, CATE: Confounder Adjusted Testing and Estimation, LFMM: Latent Factor Mixed Models. Precision is equal to 1 - false discovery rate, and F1-score is the harmonic mean of precision and power (sensitivity).

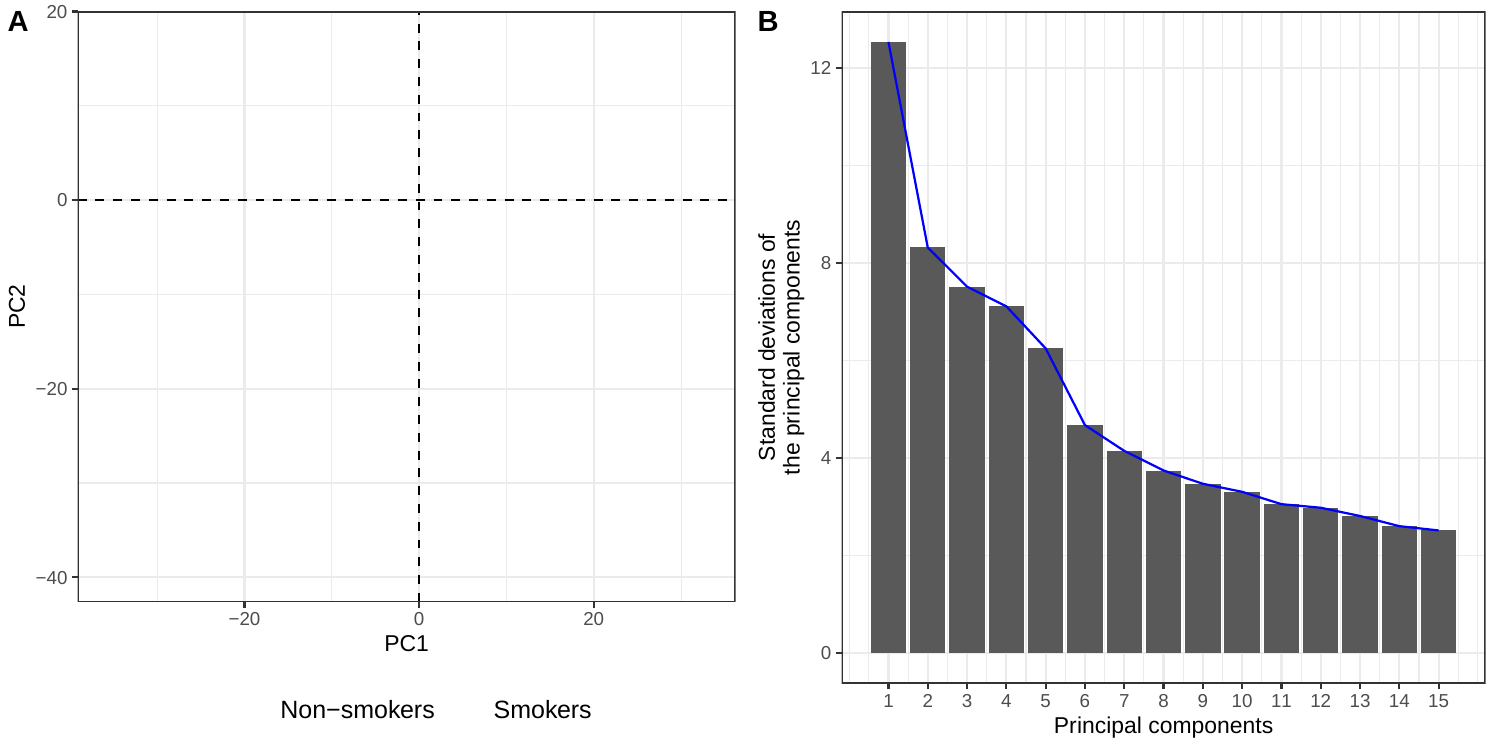

**Figure S2. Principal component analysis of placenta DNA methylation data from the EDEN mother-child cohort.** Placenta DNA methylation matrix for n = 470 women and 379,904 CpG markers. A) PC1 vs PC2, B) Scree plot of standard deviations for principal components.

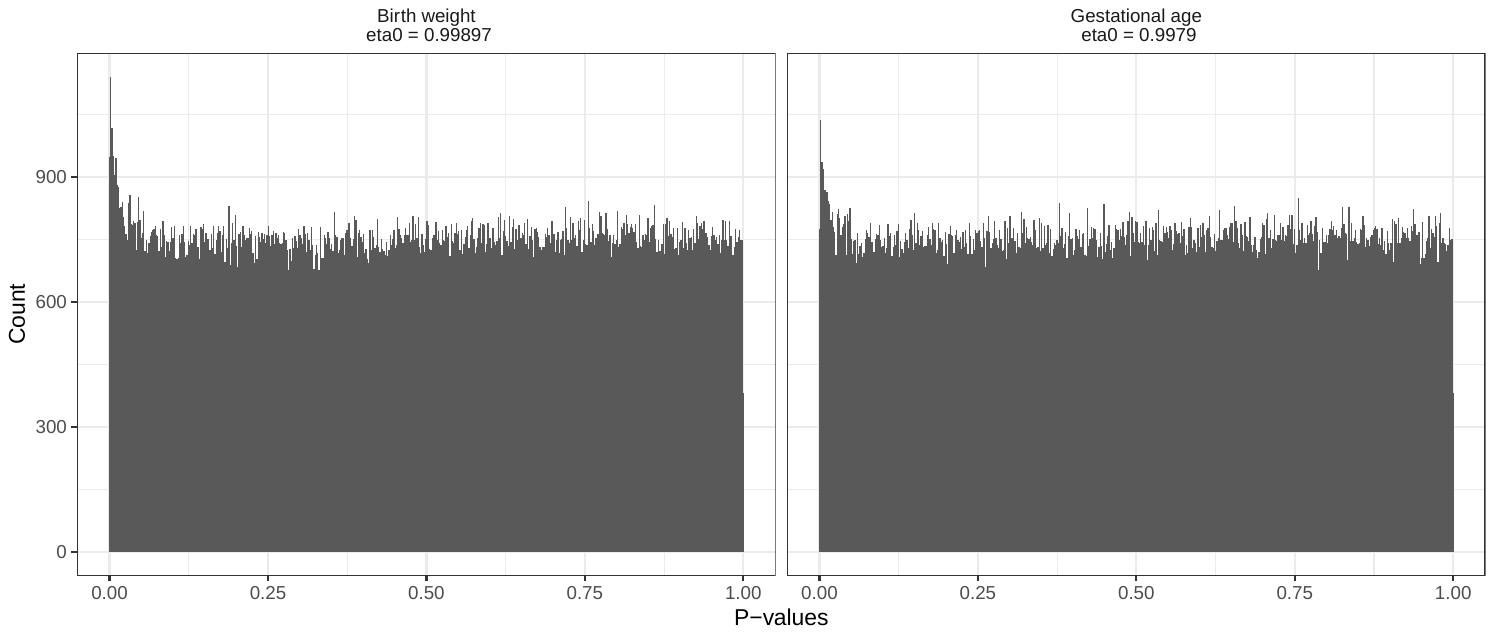

**Figure S3. Distribution of HDMAX2 *P*-values in the mediation analysis of smoking on birth weight (A) and gestational age (B) via methylation of placental DNA.**

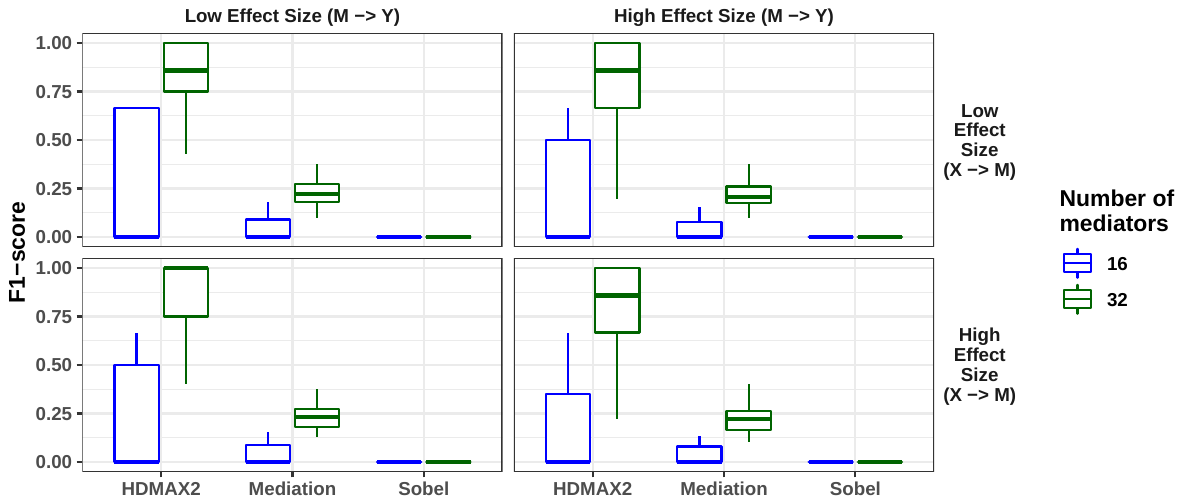

**Figure S4: Relative performances of HDMAX2 and univariate mediation methods.** F1-scores of studies analyzing the causal role of placental DNAm data in the pathway between maternal tobacco smoking and fetal growth using univariate methods or Sobel test: Morales et al 2016, and Cardenas et al 2019. They are reported according to the number of mediators (16 or 32), the effect sizes of exposure on methylation X->M (low = 0.2, high = 0.4), and the effect size of methylation on health outcome M->Y (low = 0.2, high = 0.4). Simulations of 38,000 CpGs for n=500 samples, with 9 latent factors (including 6 cell types and 3 confounding factors).

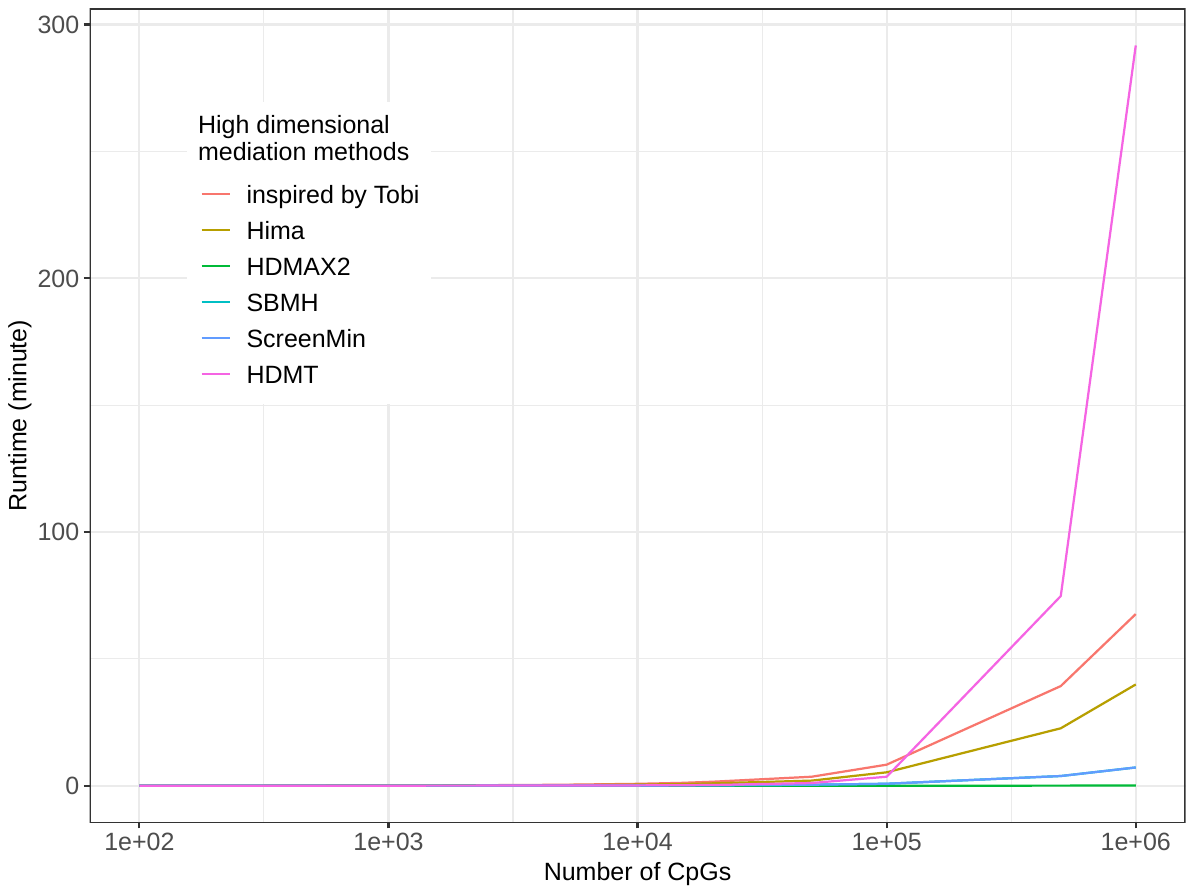

**Figure S5. Computational time for high-dimensional mediation methods according to the number of markers (CpGs).** The number of CpGs and the runtimes in minutes are shown in the log scale. The curves for the ScreenMin and SBMH methods overlay because their runtimes are equal. See the main text for the acronyms of methods.

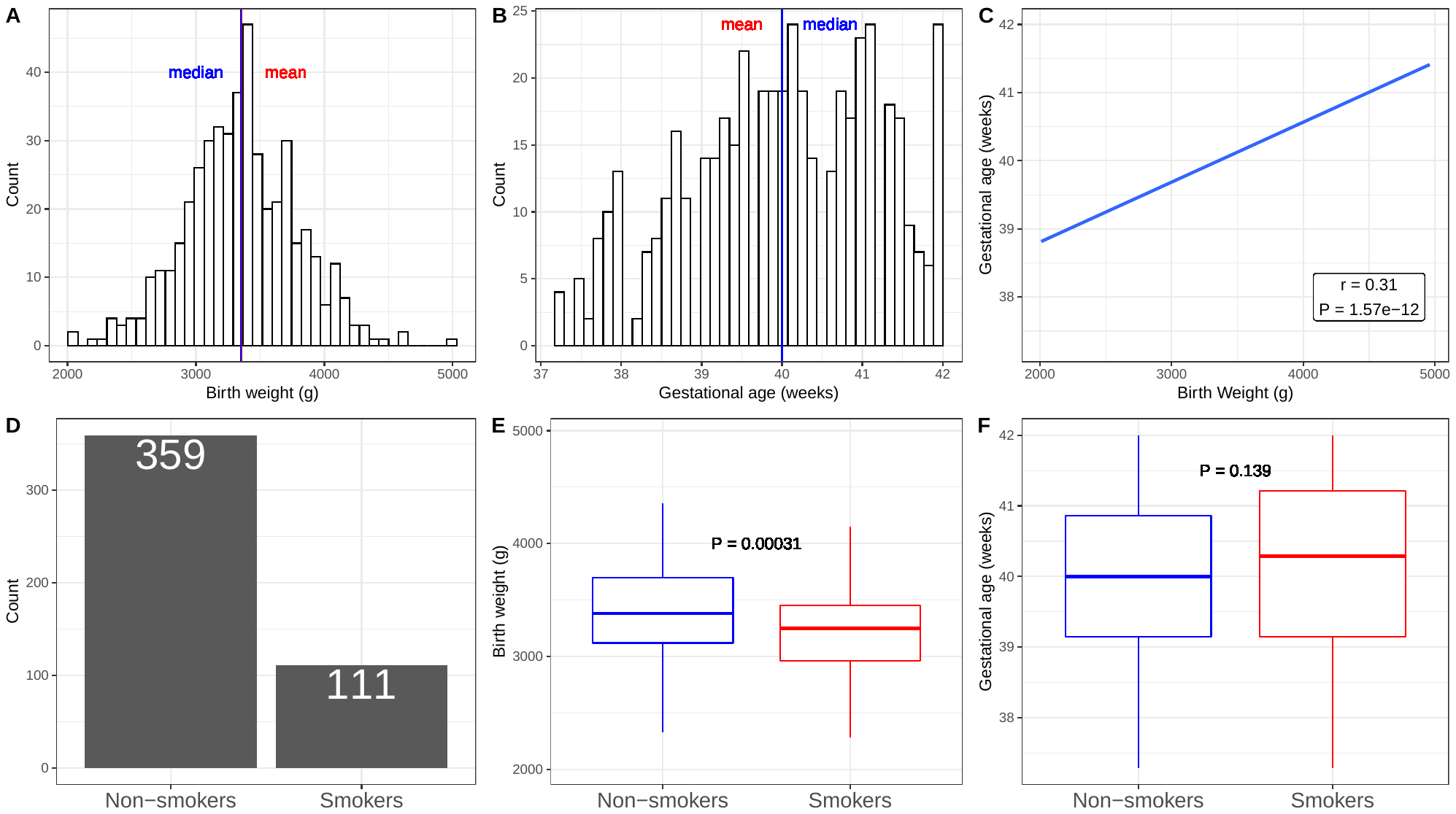

**Figure S6. Description of exposure data and health outcomes in the EDEN mother-child cohort.** A) Distribution of birth weight. B) Distribution of gestational age. C) Birth weight as a function of the gestational age. D) Distribution of smokers and non-smokers. E) Distribution of birth weight for smokers and non-smokers. F) Distribution of gestational age for smokers and non-smokers.

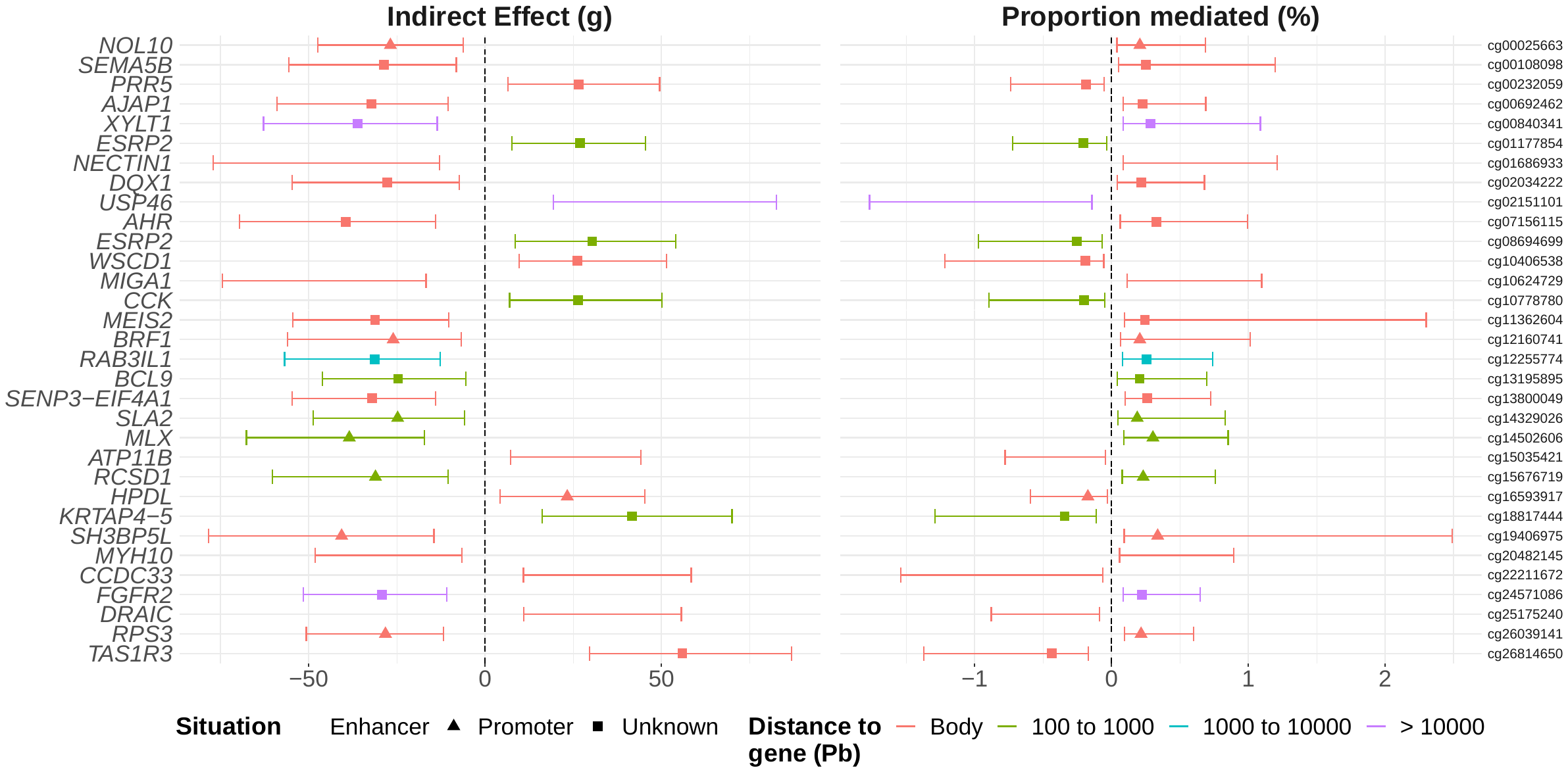

**Figure S7: high Dimensional Mediation analysis (HDMAX2) of maternal smoking on birthweight - Indirect effects for 32 CpGs.** The left panel corresponds to the estimates of indirect effects (ACME) and the right panel corresponds to the estimates of the mediated proportions. Error bars represent 95% confidence intervals for indirect effects (FDR level < 10%). The overall indirect effect of CpGs is -40.3 g (sd = 51.3).

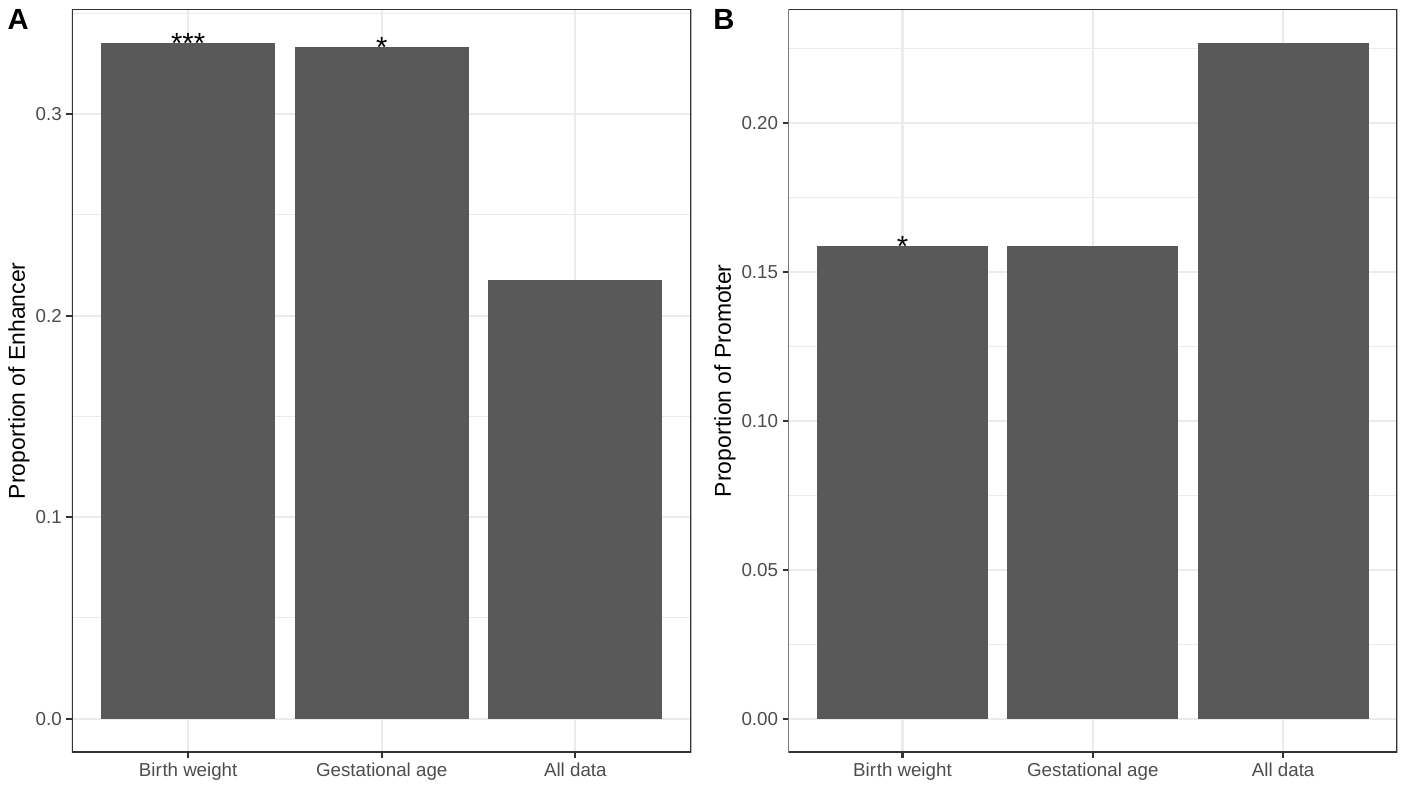

**Figure S8. Enrichment in promoter and enhancer regions.** Proportions of CpGs associated with birth weight and gestational age in enhancer and promoter regions (FDR = 20% compared to the proportions of enhancer and promoter in the full dataset (379,904 CpG). Enrichment test: *** corresponds to a significant P-value at the level P < 0.001 and * corresponds to a significant P-value at the level P < 0.05 (Fisher test).

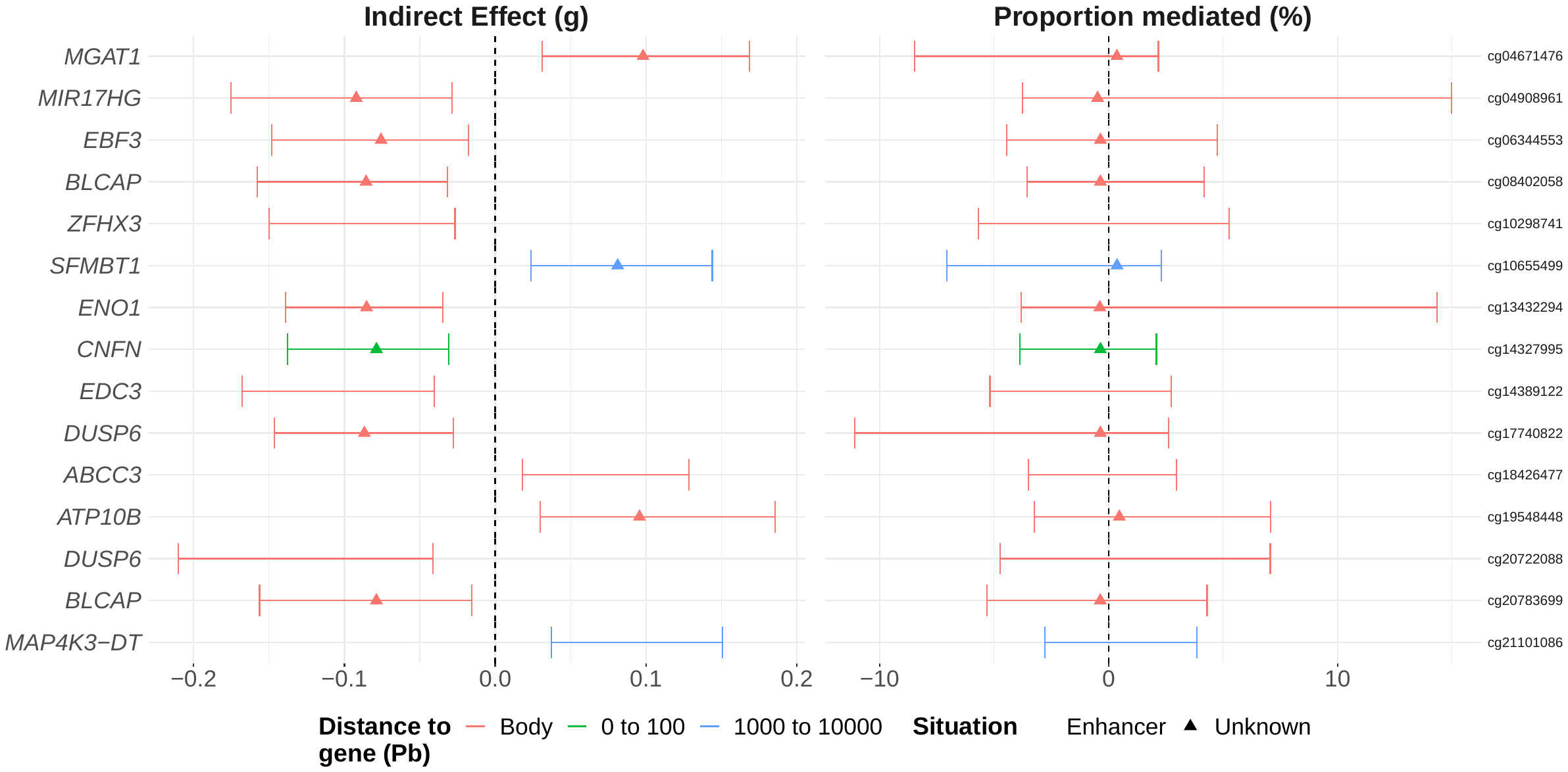

**Figure S9: High dimensional Mediation analysis (HDMAX2) of maternal smoking on gestational age - Indirect effects for 15 CpGs.** The left panel corresponds to the estimates of indirect effects (ACME) and the right panel corresponds to the estimates of the mediated proportions. Error bars represent 95% confidence intervals for indirect effects. FDR 10%. The overall indirect effect of CpGs is -0.28 weeks (sd = 0.12).

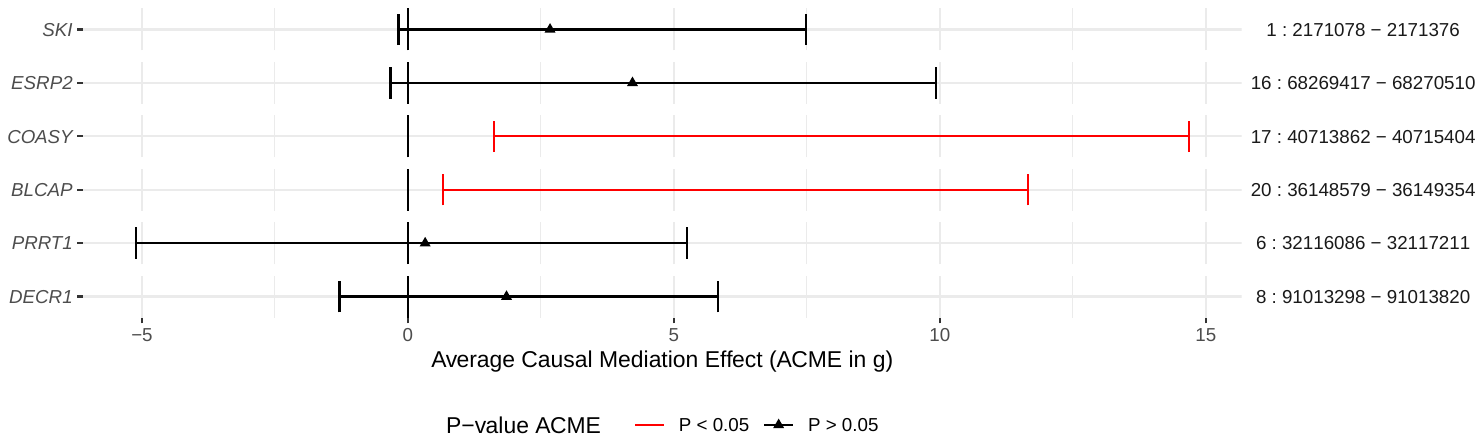

**Figure S10: Indirect effects for 6 AMRs in the mediation of GA on BW.** Indirect effects were computed for DMRs detected either as mediators of MS on BW and as mediators of MS on GA. The error bars represent 95% confidence intervals. Joint indirect effect of the 6 DMRs is 21 g (sd = 7.8).

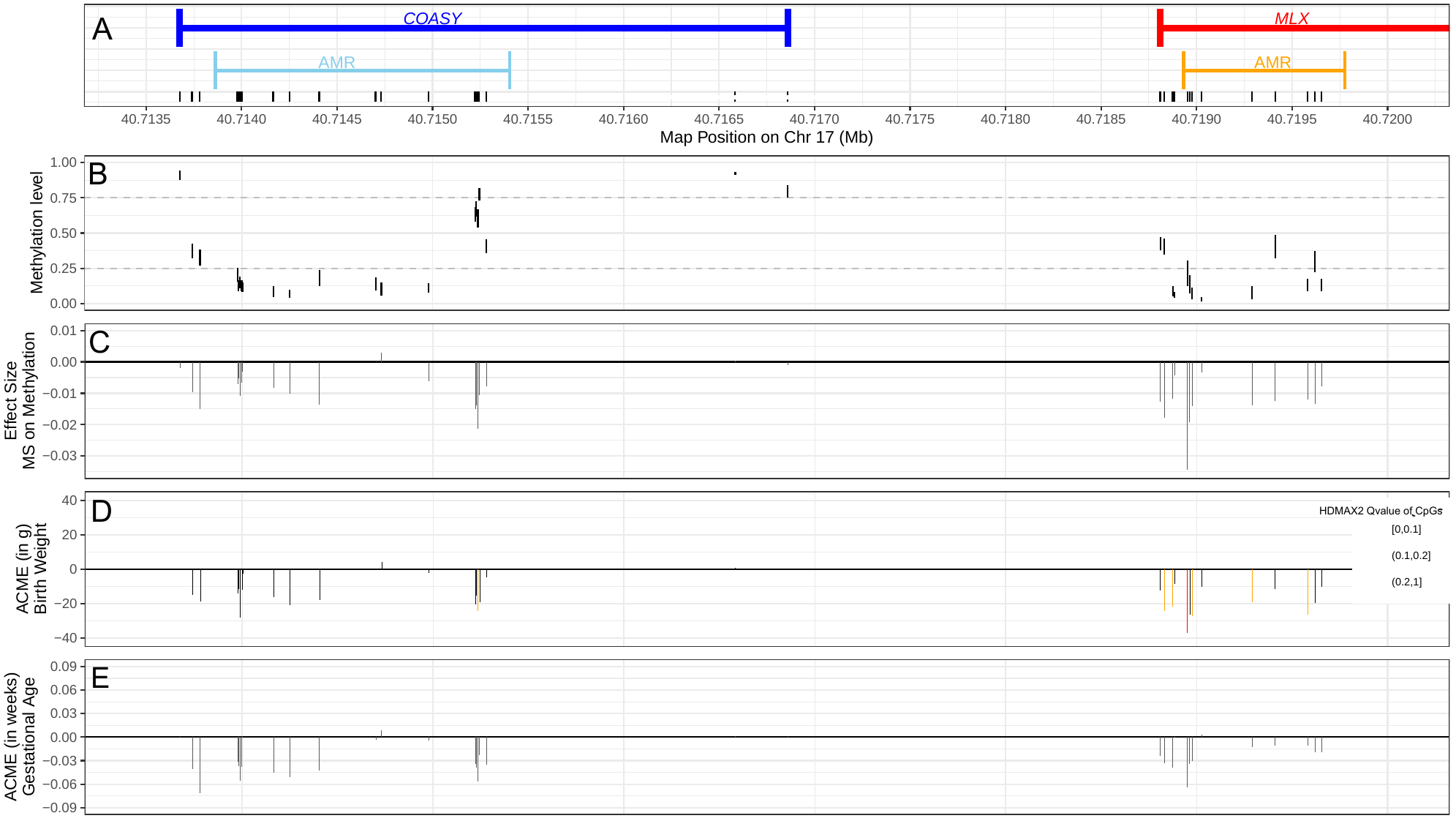

**Figure S11. AMRs in the COASY - MLX region mediating the effect of MS on BW and on GA.** A) Chromosomal positions of CpGs within AMRs, B) Mean and standard deviation of methylation levels for CpGs in AMRs showing most CpGs as being hypomethylated. C) Effect size of MS on DNAm levels. The color legend is for CpGs q-values. D) Indirect effects of MS on BW for each CpG. E) Indirect effects of MS on GA for each CpG.

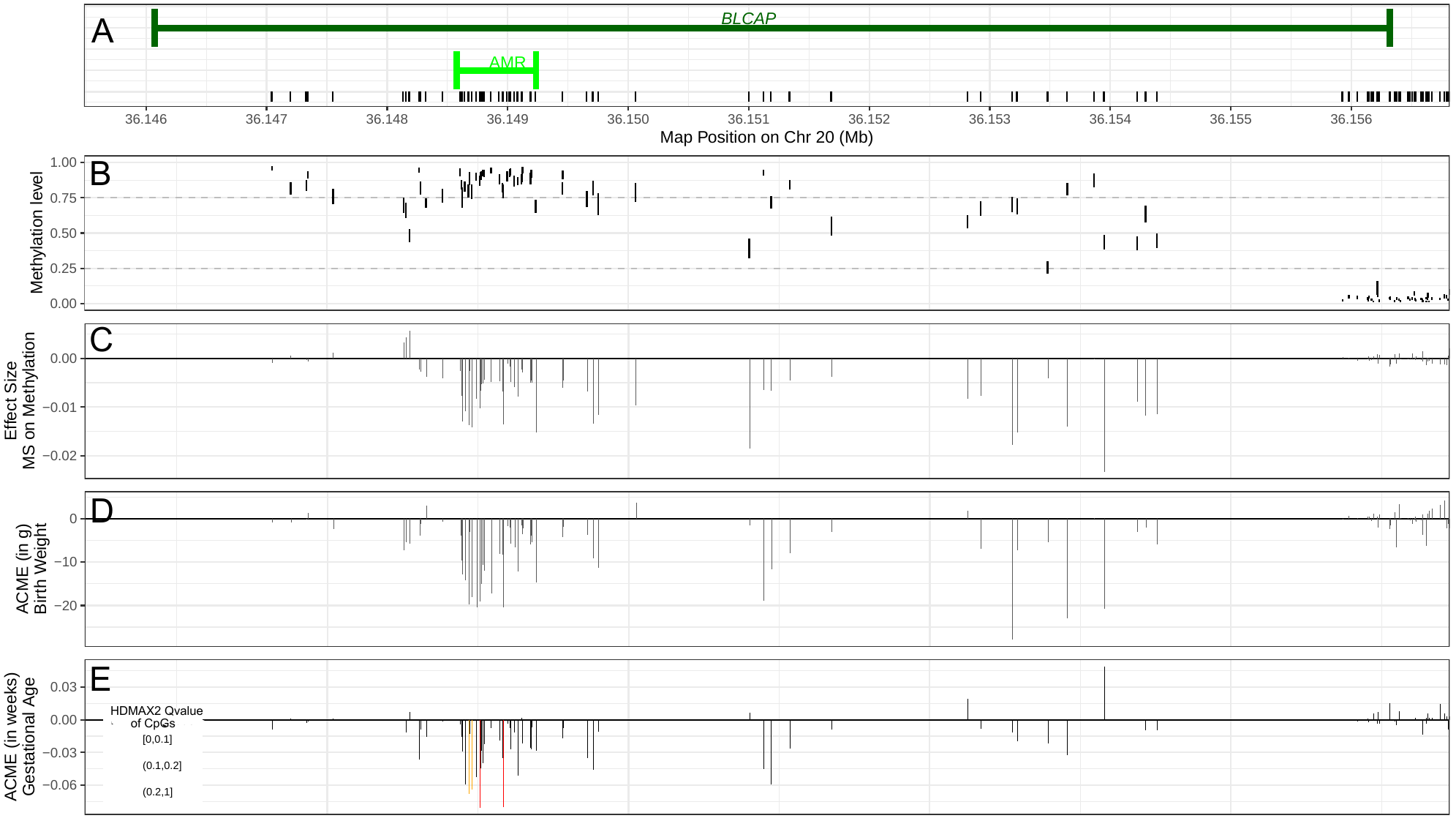

**Figure S12. AMRs in the BLCAP region mediating the effect of MS on BW and on GA.** A) Chromosomal positions of CpGs within AMRs, B) Mean and standard deviation of methylation levels for CpGs in AMRs showing most CpGs as being hypermethylated. C) Effect size of MS on DNAm levels (CpGs). D) Indirect effects of MS on BW for each CpG. E) Indirect effects of MS on GA for each CpG. The color legend is for CpGs q-values.

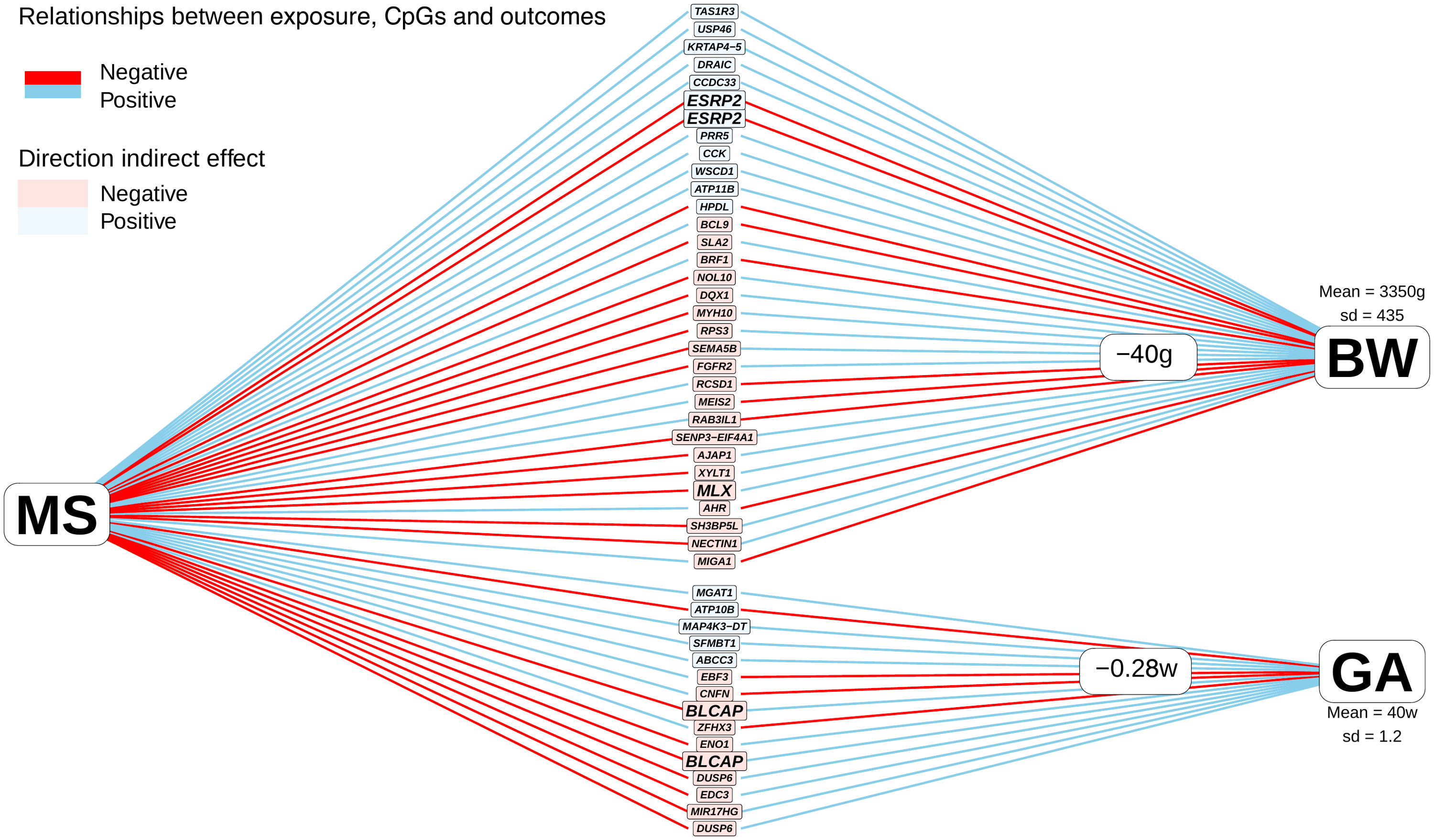
**Figure S13: Summary of CpG mediators of MS on GA and BW.** 32 CpGs mediate the relationship between MS and BW, with a total indirect effect corresponding to a decrease in BW of 40 g. 15 CpGs mediate the relationship between MS and GA, with a total indirect effect corresponding to a decrease in GA of 0.28 weeks. The color, under the genes, shows whether the indirect effect is positive or negative. The color of the segments shows whether the effect sizes, between MS and genes and between genes and outcomes, are positive or negative.

Table S1. **High Dimensional** **Mediation Analysis (HDMAX2) of maternal smoking on birth weight: CpGs found at FDR 10%.**

See excel file

Table S2. **Mediation analysis of maternal smoking on birth weight: AMRs found (FDR 5%).**

See excel file

Table S3. **High Dimensional** **Mediation Analysis (HDMAX2) of maternal smoking on gestational age: CpGs found at FDR 10%.**

See excel file

Table S4. **Mediation analysis of maternal smoking on gestational age: AMRs found (FDR 5%).**

See excel file

Table S5. **Mediation analysis of gestational age on birth weight.** AMRs common in both analyses mediating the impact of MS.

| AMR (Chr:start-end) | Gene | Indirect Effect | |
| --- | --- | --- | --- |
|  |  | beta | P-value |
| 17:40713862-40715404 | *COASY* | 6.88 | 0.00 |
| 20:36148579-36149354 | *BLCAP* | 5.12 | 0.01 |
| 16:68269417-68270510 | *ESRP2* | 4.22 | 0.12 |
| 1:2171078-2171376 | *SKI* | 2.67 | 0.10 |
| 8:91013298-91013820 | *DECR1* | 1.85 | 0.22 |
| 6:32116086-32117211 | *PRRT1* | 0.32 | 0.88 |
